# Evaluation of special educational needs and disability provision in English primary schools using administrative health and education data in the ECHILD database

**DOI:** 10.1101/2025.08.31.25334778

**Authors:** Ruth Gilbert, Jenny Saxton, Ayana Cant, Kate Lewis, Vincent Nguyen, Hayley Gains, Ania Zylbersztejn, Joachim Tan, Laura Gimeno, Jacob Matthews, Isaac Winterburn, Jugnoo Rahi, Johnny Downs, Stuart Logan, Will Farr, Lorraine Dearden, Katie Harron, Tamsin Ford, Bianca L De Stavola, the HOPE study team

## Abstract

**Introduction:** Schools worldwide balance whole-class teaching with additional provision for children with special educational needs or disability (SEND). Robust evidence on equity and effectiveness of SEND provision is essential to address growing demand and rising costs globally.

**Objectives:** We used the Education and Child Health Insights from Linked Data (ECHILD) database to examine variation in recorded SEND provision in England and its impact on health and educational outcomes in primary schools and mixed methods to understand the SEND service context.

**Methods:** We followed children from birth to age 11 years to examine social, school, and area-level factors associated with SEND provision among the general population and among children with health conditions likely to affect learning. Using target trial emulation, we estimated the impact of SEND provision on hospital admissions, school absences and attainment. We surveyed and interviewed young people, parents, and professionals and reviewed information about services to understand SEND processes and contexts.

**Results:** Of 3.8 million children born 2004 to 2013, 30% had SEND provision recorded by age 11. Health conditions are only partially associated with SEND provision, which was also related to male gender, social disadvantage, low attainment and type of school. SEND provision reduced rates of unauthorised absences but did not reduce hospital admissions or improve attainment. Mixed methods studies highlighted benefits of early, responsive support, challenges posed by limited capacity, harms caused by delayed or inadequate provision, and need for parent advocacy to access SEND provision.

**Discussion:** Weak evidence of benefits of SEND provision in causal analyses is likely to reflect unmeasured confounding, insensitive outcomes, and poor measures of the content of SEND provision in ECHILD data. SEND policies require stronger evidence from collaborative analyses across jurisdictions, based on more granular data on need, provision, confounders and outcomes, combined with experimental methods and contextual evaluation.

## Introduction

### Background

Education systems worldwide need to balance delivery of effective mainstream education with additional activities or support for children who experience difficulties with learning compared to their peers.[1] In this study of the English context, we refer to such additional support as special educational needs or disability (SEND) provision (see Appendix 1, Table S1).[2][3] Typically, SEND provision involves a wide range of types and intensity of activities, from continuous one-to-one support to occasional group work, across various types of needs. Activities change as the child responds and develops.[4] SEND provision also includes collaboration between multi-sector professionals and families, all within a complex, publicly funded system.[5]

Children likely to need SEND provision include those with disability or chronic health conditions that affect learning. In many countries, children with behaviour problems, speech or language difficulties, low school attainment, social disadvantage, and those who speak a different language are also assigned SEND provision.[1] Countries worldwide report rising demand and increasing costs of SEND provision, reflecting improved survival among children with medical conditions, increasing recognition of neurodevelopmental disorders, and global efforts to expand educational access and provide inclusive education for all.[6] [7] [8]

### Existing evidence

Evidence for effective SEND provision is mixed. On the one hand, an international systematic review of 467 comparative studies of children with known additional needs (including 297 randomised controlled trials; RCTs), reported consistent beneficial effects of different types and durations of SEND activities compared with service as usual or alternative education activities.[5][9] On average, these benefits equate to 5 to 6 months of improved learning in maths, reading, or related skills. However, most of these studies focused on targeted activities for well-defined learning difficulties (e.g. 40% were for dyslexia), rather than evaluating SEND provision across the breadth of usual practice. In addition, the impact of these targeted activities, measured under research conditions, may not be generalisable to SEND processes as typically delivered as part of a complexity of activities, relationships and services in different school and classroom contexts.

Such a complex process is hard to evaluate. So far, limited evidence of benefit has been reported, by numerous observational studies over the past 25 years. As expected, descriptive studies found that children assigned SEND provision had worse outcomes than peers, due to confounding by higher underlying health and education needs.[10][11][12] However, 49 studies that attempted to account for confounders using quasi- or natural experimental designs, also found no difference or worse outcomes for children assigned SEND provision. [12][13][14][15][16][17][18] Similarly adverse findings were reported in an evaluation of teaching assistants in England, a group who are typically involved in SEND provision.[19]

Recent advances in causal methods are presented as a framework for using natural experiments, sometimes called quasi-experimental methods, to evaluate population health interventions.[22] The report refers to a range of methods to achieve comparability (or exchangeability) between those with and without intervention. Various methods can be used to account for measured confounding, including those within the target trial emulation framework (TTE).[21] Methods often used to address unmeasured confounding include difference-in-difference and instrumental variable techniques A meta-analysis of 44 U.S. studies that tried to minimise confounding found eight studies that addressed the bias arising from unmeasured confounding.[15] These studies used policy changes,[23][24] or changes within children’s SEND status over time,[25] [26] [27] or differential SEND thresholds across racial groups in natural experiment approaches.[28] All eight found effects of SEND provision: five reported beneficial impacts, while three reported harmful effects for certain subgroups. For example, two studies found worse outcomes for children who were just below the threshold for SEND provision, who received provision due to policy changes.[28][29] These effects for marginal groups might not be generalisable to all children who need SEND provision.

In summary, uncertainty remains about the benefits of SEND provision as delivered in routine practice globally, as well as in England. At the same time, there is strong evidence that targeted activities, delivered as a key element of the SEND process, are effective, albeit often for narrowly defined needs and outcomes.[9] Policy makers need robust evidence on what types of SEND provision work, for whom, and under what conditions, and on the effectiveness of the overall SEND process, to deliver effective support while containing costs. Applying robust study designs to linked administrative health and education data provides a unique opportunity to fill these evidence gaps.

### SEND policy in England

Despite the uncertain evidence, major policy changes have occurred to funding and delivery of SEND provision over recent decades, which offer opportunities for natural experiment designs that address unmeasured confounding.[2][31] Details of these changes are summarised in Appendix 1 (Box 1).[32] Further reforms to the SEND provision system will be published by the Department for Education (DfE) in Autumn 2025.[33] [34]

### Purpose of this paper

This paper presents an overview of an evaluation of the impact of SEND provision in England and discusses implications for SEND policy and for future, international research using linked administrative health and education data. The Health and education Outcomes of young People throughout Education (HOPE) study is the first study to use the newly linked health and education administrative data from the ECHILD database to evaluate the impact of SEND provision in England.[35] Lessons learned are relevant to how policy makers, funders and researchers make best use of ECHILD and similar datasets for future evaluations. Further details can be found in the protocol and 29 other papers listed in Appendix 4.[32]

## Study design and methods

### Aims and research questions

The HOPE study aimed to examine variation in SEND provision in English primary schools and assess its impact on children’s health and educational outcomes.[32] We addressed four main research questions (Figure 1) in consultation with young people, parent-carers and professionals throughout the study (Appendix 2):

**1)** Which health conditions (phenotypes) are associated with outcomes that might be improved by SEND provision?
**2) What factors influence who is assigned SEND provision, when and where?**

- **2a:** Who is assigned SEND provision?
- **2b:** When is SEND provision assigned?
- **2c:** Does where the child lives and goes to school influence SEND provision?
- **2d:** Was SEND provision affected by the 2014 education reforms?
**3) What is the impact of SEND provision on health and education outcomes?**
**4) What do service experiences and policies tell us about:**

- **4a:** initial SEND identification at primary school?
- **4b:** formal SEND assessments and the development of provision plans?
- **4c:** the implementation of SEND-related provision?
- **4d:** outcomes for children and families from SEND processes and activities?
- **4e:** local service capability for implementing SEND policy?
- **4f:** wider influences on local SEND service capability and functioning?

**Figure 1.**
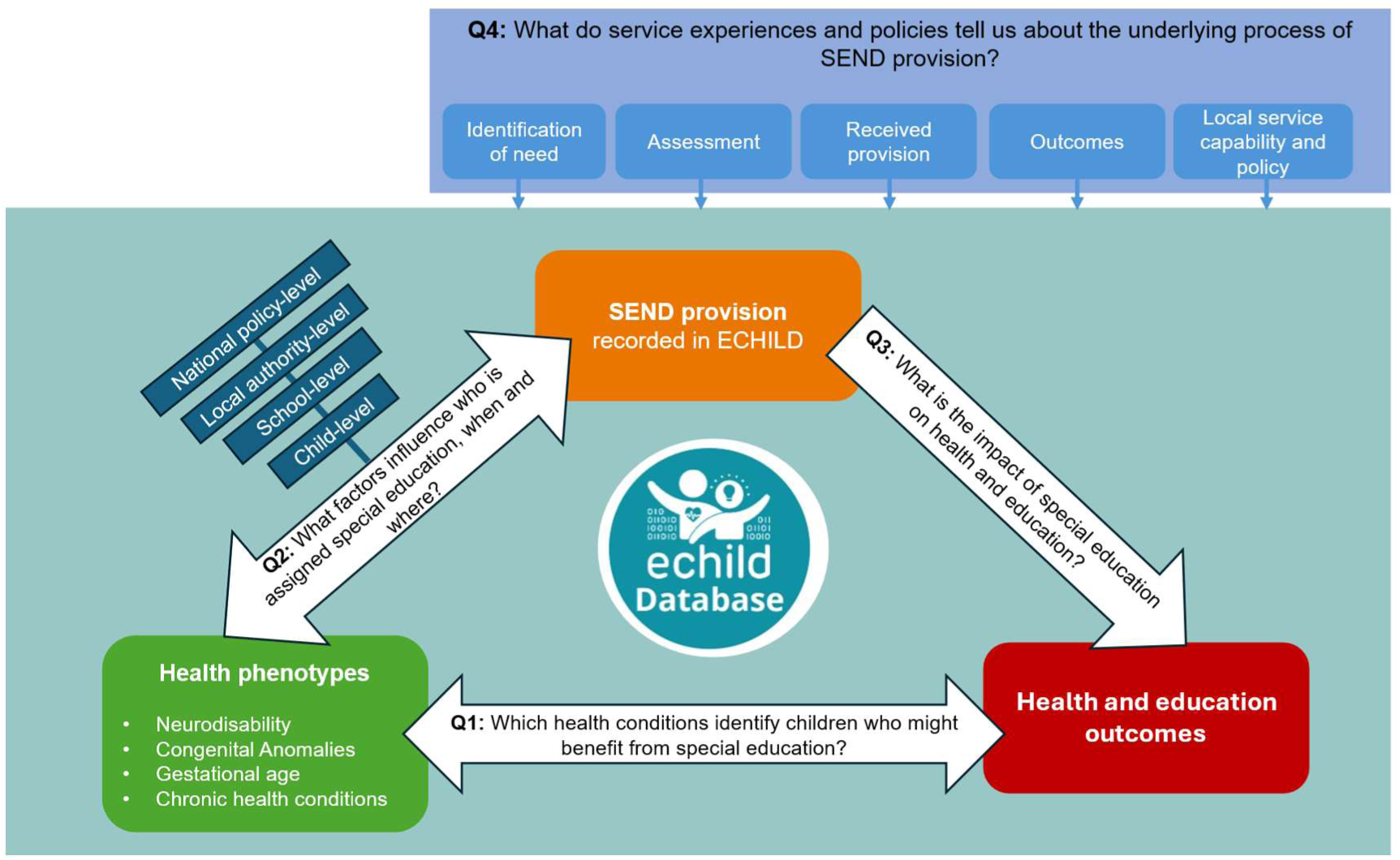
Diagram of research questions addressed by the HOPE study.

### The ECHILD database

The HOPE study used administrative data from the ECHILD database for all analyses related to questions 1–3 (Appendix 1, Table S1 for details and key variables).[35] We defined birth cohorts of singleton children born in NHS hospitals in England. Children were followed from birth through to the end of primary school (Year 6, age 10/11) or August 2019 (whichever occurred first). Ending follow up in 2019 avoided the COVID pandemic (from March 2020), when hospital contacts as well as school absence and attainment changed or were not consistently recorded.[32][36] Different sub-cohorts were extracted to address the questions 1 to 3 (Appendix 3).

### Question 1: Which health conditions (phenotypes) are associated with outcomes that could be improved by SEND provision?

The first step of the HOPE study involved developing health phenotypes to proxy groups of children likely to need SEND provision. This step is critical, as education data captures only the assignment of SEND provision but no indicators for the underlying need for additional support. Using external evidence, clinical input and insights from ECHILD data regarding the reliability of diagnostic coding in administrative data, we chose three groups of health phenotypes which capture populations with different levels of need for SEND provision: neurodisability,[37] congenital anomalies,[38] and preterm or early term births, defined by week of gestational age.[39] [40] These phenotypes were mainly recorded before school entry and, on average, were expected to have mild to moderate (congenital malformations) or moderate to high (neurodisability) need for SEND provision or a gradient of need by week of gestational age at birth. We described outcomes for these phenotypes and between subgroups within each phenotype. Combinations of neurodisability and other health phenotypes were described for children with Down syndrome (Appendix 3, extended cohort 1; Appendix 4, paper 10).

We assessed outcomes for children with these phenotypes and unaffected peers, from school entry to Year 6 (age 11) using cohorts 1-5 (Appendix 4, papers 2-9).[40][41][42][43] Outcomes included: mortality before school start, planned and unplanned hospital admissions, unauthorised school absences, and whether a child reached the expected level of development, based on school test scores at ages 5, 7 and 11 (Appendix 1, Table S1)

### Question 2: What factors influence who is assigned SEND provision, when, and where?

Question 2 involves descriptive analyses to understand different ways in which SEND provision varies. Findings contributed evidence on variation in SEND services and were used to define analyses and comparison groups and to account for confounding in the causal analyses (question 3).

### 2a) Who is assigned SEND provision?

We described the proportion of children with any SEND provision (SEN Support and/or an Education, health and care plan - EHCP) in Year 1 and up to Year 6 for each of the three health phenotypes (neurodisability, cohort 2; congenital anomalies, cohort 3,5; gestational age, cohort 4) and their subgroups, and for unaffected peers (Appendix 3).[44] [38] [40]

In separate analyses of the whole population of children we described the proportions of children who were assigned SEN Support, EHCP, or any SEND provision in Year 1 (age 5/6) – the first full year of compulsory education - according to exposures recorded in education or health data before Year 1 (Appendix 3, cohort 6).[45] These exposures included proxy health needs (any recorded chronic health condition, preterm birth <37 weeks of gestation), gender, month of birth to denote youngest in class, social disadvantage (recorded free school meal eligibility – indicating receipt of welfare benefits), living in the 40% most deprived neighbourhoods, being born to a young mother (aged <20 years), and school readiness measured at age 5 by i) a standardised mean score or ii) not attaining a good level of development in the Early Years Foundation Stage Profile (EYFSP) (Appendix Table S1).

### 2b) When is SEND provision assigned?

We analysed the whole population of children born between September 2006 and August 2008 (cohort 6) to assess the stage at entry into state education. By state education, we mean state-provided Nursery (school-based or local authority (LA) provided at age 2-4) or Reception class (age 4/5) in a state-funded primary school. By stage, we mean first entry into state education either in Nursery (N1 age 2 or N2 age 3), or Reception, or Year 1. We plotted the percentage of children assigned SEN Support or EHCP during state-funded hours in any Nursery setting (including school-based, LA-provided, private or voluntary), or in state primary school from Reception class to Year 6, according to stage at entry to state education (see Appendix 4, paper 15).[46] Eligibility for state-funded hours in nursery is given in Appendix 1[45].

To understand how timing of SEND provision varied for children with high need for SEND provision, we conducted separate analyses of a cohort of children with cerebral palsy recorded in hospital admission records before entering state education (Appendix 3, cohort 8; Appendix 4, paper 12). We used a staggered cohort design to account for early entry into state school-based Nursery (age 2 or 3), or Reception class (age 4/5). At each stage at entry to education, and combination of sociodemographic variables (gender, neighbourhood deprivation and free school meal status), we estimated the average cumulative probability of any SEND or EHCP provision, by year from Nursery to Year 6, using inverse probability weighted logistic regression.[47]

### 2c) Does where the child lives and goes to school influence SEND provision?

Schools and LAs make decisions about SEND provision.[4] We first assessed whether the type of school governance in Year 1 influenced SEND provision in the same year by analysing the whole population of children born in cohort 6 (Appendix 3). School governance affects admissions policies, teaching about faith, and staff employment (Appendix 1, Table S1). A shift to more autonomous schools, not controlled by the LA, has been progressing since 2007.[48] We plotted the observed probabilities for any SEND, SEN Support or EHCP provision by school type. We then estimated equalised probabilities by controlling for (and then averaging over): the stage at entry to state education (Nursery, Reception, or Year 1), chronic health conditions, preterm birth, social disadvantage, demographic factors prior to stage at education entry and the EYFSP score taken at age 5 (Appendix 4,paper 15).[45]

In separate analyses, we also assessed whether the LA of residence, which is responsible for funding and assigning EHCPs, influenced the probability of a child being assigned SEND provision using three subgroups of children born in 2003 to 2014 (Appendix 3, cohort 7).[49] [50] Subgroups were defined by gestational age groups expected to have diminishing need for SEND provision: late preterm (34–36 weeks), early term (37–38 weeks), and full term (39–41 weeks).[49] We quantified variation between LAs in the probability of receiving SEN Support, EHCP or any SEND provision in Year 1, by estimating the intraclass correlation coefficient (ICC) at each stage of multilevel models that controlled for child level health, social and demographic factors, EYFSP score age 5, school type (mainstream or special) and school governance (Appendix 1, Table S1). We also controlled for LA characteristics, including total number of pupils, proportion eligible for free school meals, the modal deprivation quintile of children and percentage on a child protection plan (Appendix 4, paper 14).[49]

### 2d) Was SEND provision affected by the 2014 education reforms?

The Children and Families Act (2014),[2] enacted through the SEND Code of Practice (2015),[4] introduced changes to SEND provision. We analysed children with major congenital anomalies and their unaffected peers who were born in 2003 to 2014 (Cohort 5), We assessed evidence of sudden change in the probability of SEN Support or EHCP provision in Year 1 (age 5/6) between periods before (2009-2014) and after the reforms (2014-2019; Appendix 3, Cohort 5; Appendix 4, paper 9).[38]

### Question 3: What is the impact of SEND provision on health and education outcomes?

We planned to use two quantitative approaches to evaluate the impact of SEND provision on health and education. Both approaches are within the natural experimental framework; both use data emerging from an event or process associated with the introduction, delivery or withdrawal of an intervention to evaluate the impact of the intervention on outcomes.[21] The first approach involved assessing whether observed changes in policy (Question 2d above), or differences in practices between schools or local authorities (Question 2c), introduced sufficient exogenous variation to enable the use of instrumental variable methods for the estimation of the impact of the new policies (Appendix 1). These methods can account for unmeasured confounding.[20][21]

The second approach used the target trial emulation (TTE) framework to define causal questions that could replicate an ideal RCT with observational data.[51] The framework requires defining the causal contrasts of interest, the exposure (SEND provision), the relevant outcomes and confounders. Principled methods to deal with measured confounding, such as propensity score-based and g-methods are then adopted. By making all elements of the causal question explicit, the TTE framework helps to minimise biases, for example, due to mis-alignment of study entry, eligibility and exposure assessment, or to incorrect assignment of later exposure status to an earlier period.[52] The TTE framework also guides the choice of methods for estimating impact, including evaluating whether no unmeasured confounding (or conditional exchangeability) can be assumed.[51]

Using the TTE framework, we evaluated the impact of SEND Support compared with no support, EHCP with SEN Support, and EHCP with no support, in Year 1 (age 5/6) of school. We analysed two sub-phenotypes of children: those with cleft lip with palate and children with cerebral palsy (Appendix 3, cohorts 9,10; Appendix 4, papers 16,18).[53][54] We iteratively refined these phenotypes to obtain groups that were relatively homogeneous, by excluding children with anomalies affecting other body systems. Outcomes were unplanned hospital admissions, unauthorised school absence in Years 2-6 and attainment in mathematics in Year 2 (age 6/7) and Year 6 (age 11). For the cerebral palsy group we also looked at total absences. We hypothesised that SEND provision could improve participation in social and educational activities, and thereby reduce stress, promote wellbeing, social competence, behaviour and communication, and parent-child responses to developmental needs – all factors that could influence unplanned hospital admissions through mental or physical health or stress-related presentations, or injury. School absence is also influenced by these factors.[55] [56] [57] [58]

We chose cleft lip with palate as a phenotype likely to reflect mild to moderate need for SEND provision, because average differences in education and health outcomes between affected children and peers are small.[59] In addition, there are clear differences in outcomes according to the type of anomaly (isolated unilateral or bilateral cleft lip or cleft lip with palate), and whether other body systems are affected.[59][60][61]

The cerebral palsy phenotype was chosen to proxy high need for EHCP provision using specific codes for cerebral palsy, with broader criteria used in a sensitivity analysis. We compared SEN Support to none, and EHCP to SEN Support in Year 1.[54]

For both sets of analyses, estimation of causal effects required evidence of no unmeasured confounding (also termed conditional exchangeability) and positivity (overlapping propensity to be assigned to each level of exposure; see Appendix 1). In each case there was insufficient overlap in propensity scores to compare EHCP with no provision and thus this comparison was not feasible (see Appendix 4, paper 18).[53] [54]

### Question 4: What do service experiences and policies tell us about the underlying process of SEND provision?

We used mixed methods to understand experiences of the SEND process from the perspectives of young people and parent-carers using the service and from professionals contributing to SEND provision in some way.[32]

We examined three core stages of the SEND process: noticing the need, SEND assessment and plans, and provision of support. We explored wider national influences such as service capacity, relevant outcomes for the child, their sibling and parents, and wider context (Figure 2). Ten mixed methods sub-studies were used to answer questions 4a-f. These included qualitative studies (interviews and focus groups), online surveys (with children, young people, parents/carers, and professionals) and document reviews of Ombudsman complaints, Ofsted/CQC inspection reports, Local Offer (LO) websites and grey and peer-reviewed literature on variation in SEND processes and provision at LA and multi-academy trust levels (Appendix 1).

**Figure 2.**
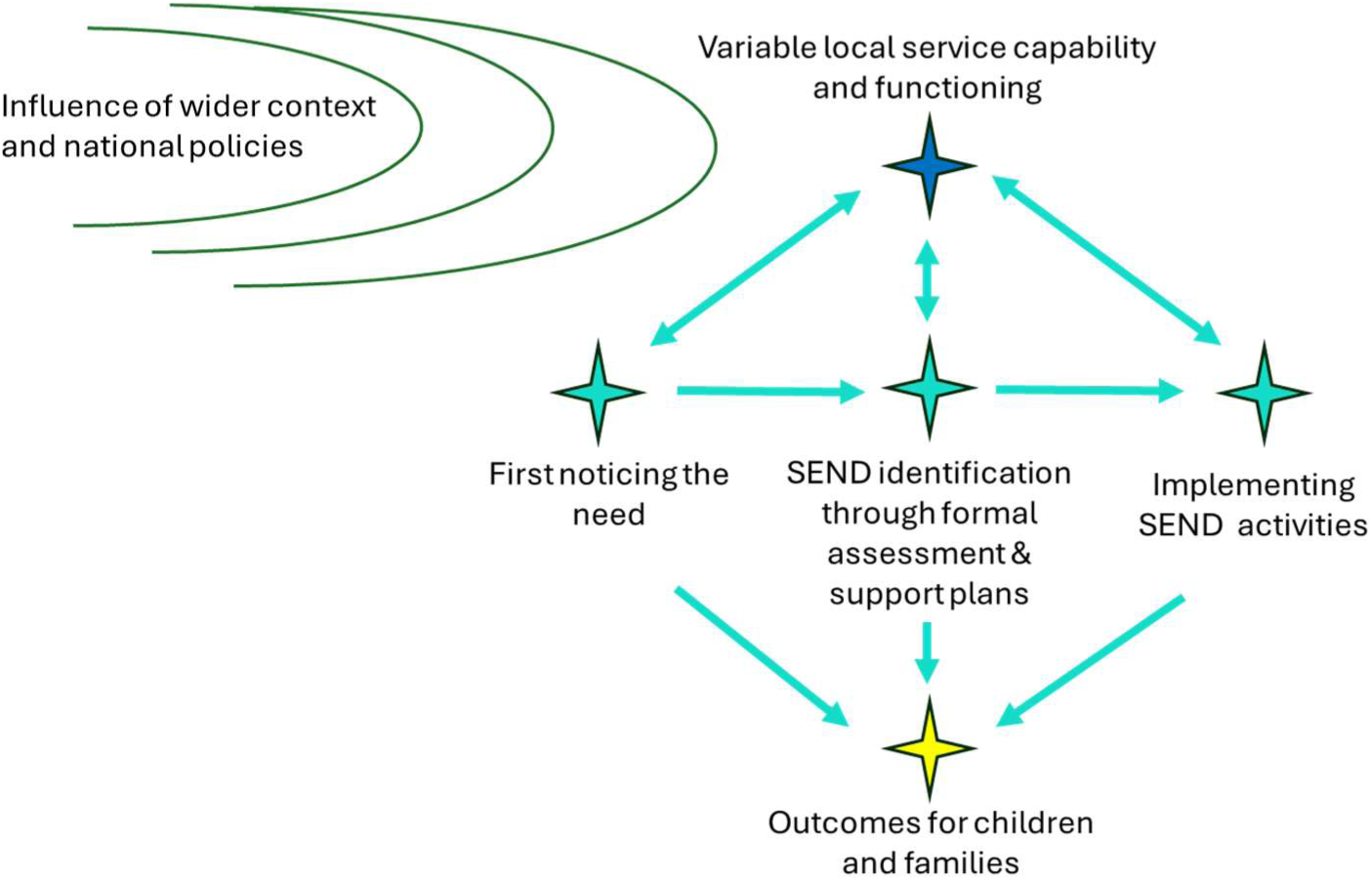
Experiences of the SEND process, outcomes, and wider influences on local service capability to deliver services.

Appendix 5 summarises methods and key findings from each of the 10 sub-studies mapped to research questions 4 a to f or see Appendix 4, papers 21-30 for further details.

## Results

### Question 1: Which health conditions (phenotypes) are associated with outcomes that might be improved by SEND provision?

Children in each of the broad phenotype groups had more unplanned admissions, more frequent absences and lower attainment than their unaffected peers. As expected, there was substantial variation between phenotype subgroups.[44] We illustrate these patterns by summarising results for children with neurodisability. Of 3.58 million children, 2.4% had a hospital-recorded neurodisability before age 5, rising to 3.6% by age 11 (Appendix 3, cohort 1 and 2; Appendix 4 paper 2).[44] These children were admitted to hospital five to seven times more often than their peers and accounted for 15% of all hospital bed-days during primary school (for the 2.96 million children who had linked education records in primary school in cohort 2).[62] We also found 60% higher school absence rates and consistently lower attainment during primary school among children with neurodisability (Figure 3)(Appendix 4, papers 3,5).[62][63] At age 5, only 30% reached a ‘good level of development’ (GLD), compared with 58% of peers, and fewer than half met expected levels at ages 7 and 11 (Figure 3a).[63] By the end of primary school (Figure 3a, key stage 2, age 11), over a third of children with neurodisability were not assessed in national attainment tests, contrasting with 6.4% of unaffected peers enrolled in school (see key stage tests in Appendix 1).[63] Outcomes varied significantly by neurodisability type and in each phenotype subgroup (Figure 3b).[62] [63]

**Figure 3.**
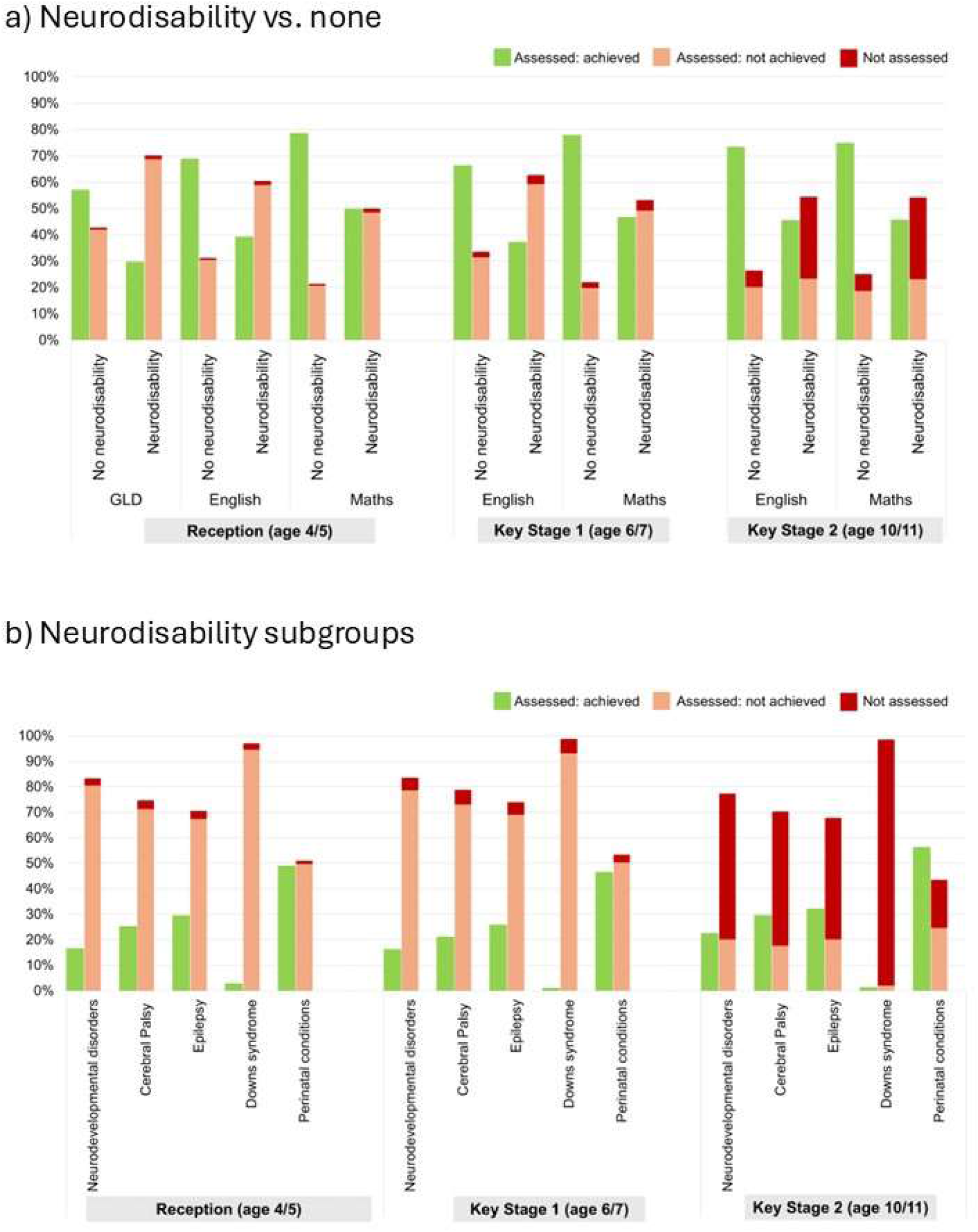
Proportion of children with and without hospital-recorded neurodisability by age 5 (and subgroups) achieving nationally expected levels across primary school assessments.

Children with congenital anomalies comprised 3.5% of 2.35 million children entering primary school (Cohort 3). Attainment varied between subgroups, with the lowest average attainment through primary school occurring among children with chromosomal or neurological congenital anomalies (Appendix 4; papers 5,7).[63][64] Average differences between children with and without congenital anomalies were small to moderate.[64] We found lower attainment across all anomaly subgroups for boys than girls.[64] Separate analyses of the whole population (Cohort 4) revealed lower attainment for each week of gestational age at birth before and after 40 weeks of gestation (Appendix 4, paper 11).[40]

#### Question 1: Key findings

- Children with neurodisability, congenital anomalies, or preterm birth had consistently more unplanned hospital admissions, more frequent school absences and lower attainment at age 7 and 11 compared to unaffected peers.
- These outcomes were on average worse for children with neurodisability than for children with congenital anomalies or those born preterm.
- Outcomes varied substantially between specific condition subgroups within each phenotype, and by week of gestational age at birth.

### Question 2: What factors affect who is assigned SEND provision, when, and where? 2a. Who is assigned SEND provision?

Our analyses of children with neurodisability (cohort 2), congenital malformations (cohort 5) and all children by week of gestational age at birth (cohort 4), showed that assignment of SEND provision was consistently higher for children with chronic health conditions or born preterm than their unaffected peers.[38][44] Those with neurodisability had the highest proportions with SEND provision: three-quarters (76%) had any SEND provision recorded ever during primary school and 40% had an EHCP.[44] The proportion of children with SEND provision varied substantially between different condition subgroups within the neurodisability and congenital anomaly phenotypes and by week of gestational age at birth (Figure 5; Appendix 4, papers 2,9,11).[38] [40][44]

**Figure 5.**
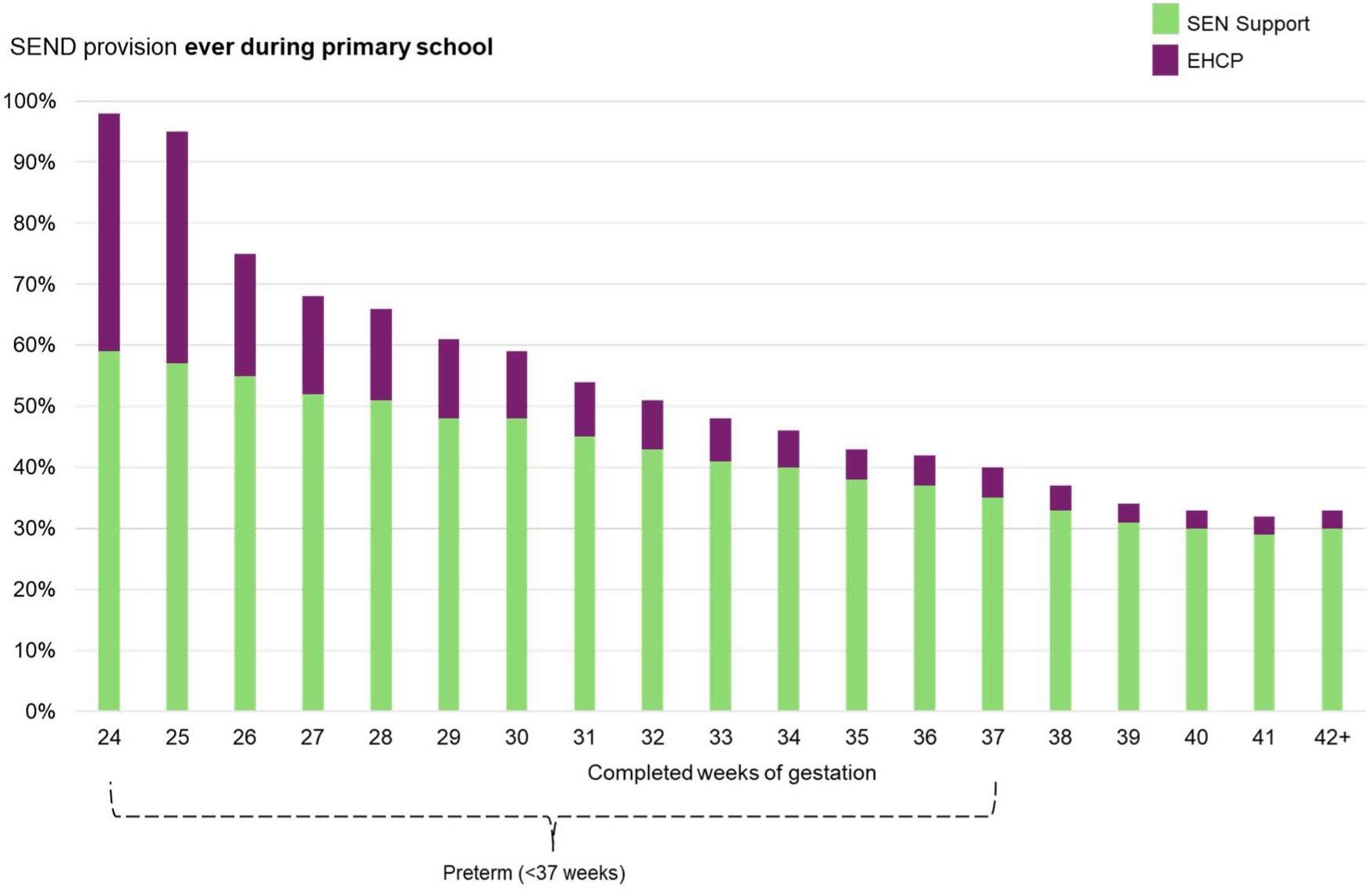
Proportion of children by week of gestation assigned SEN Support or an EHCP ever during primary school (Cohort 3, born 2003/04 to 2007/08).

In analyses of the whole population of children (Cohort 6), chronic health conditions recorded in hospital records were strongly associated with SEND provision. Nearly 1 in 5 (18.1%) of children had one or more chronic health condition recorded, of whom 26.6% had any SEND provision and 5.9% an EHCP in Year 1. Equivalent percentages for those without a chronic health condition were 13.4% and 0.7%. However, children with chronic health conditions accounted for only 30.5% of all children with any SEND provision and 63.9% of those with an EHCP in Year 1.

Many children without recorded chronic health conditions were assigned SEND provision, including EHCP, reflecting the importance of other factors affecting learning difficulties or health problems not requiring hospital admission.[45] SEND provision was more common among boys, those who were youngest in class (i.e. summer-born children), and those from disadvantaged backgrounds as measured by free school meal eligibility, young motherhood, and residence in the 40% most deprived areas (Appendix 4, paper 15).[45] However, associations between EHCP assignment and social disadvantage were weaker and less consistent, although the association with boys remained strong.[45]

### 2b. When is SEND provision assigned?

Using longitudinal data for the whole population of children born between September 2006 and August 2008 (Cohort 6) we described the stage at entry to state education: two in five children (42%) started state-provided Nursery at ages 2 to 3 and just over half (57.6%) started in Reception (Appendix 4, paper 15).[45] Children starting in Nursery had higher levels of chronic health conditions and social disadvantage, consistent with the socioeconomic criteria for accessing free nursery places at ages 2 to 3.[45][65]

The cumulative percentage of children assigned SEN Support or EHCP provision up to Year 6 was highest for those starting state education in Nursery and for the few (0.4%) entering in Year 1. It was lowest for those starting in Reception (Figure 6; Year 1 entrants not shown). The percentage assigned SEN Support increased gradually, peaked in Year 2, and then levelled off. In contrast, the percentage assigned EHCP provision remained very low (<5%) and rose more slowly throughout primary school.[45]

**Figure 6.**
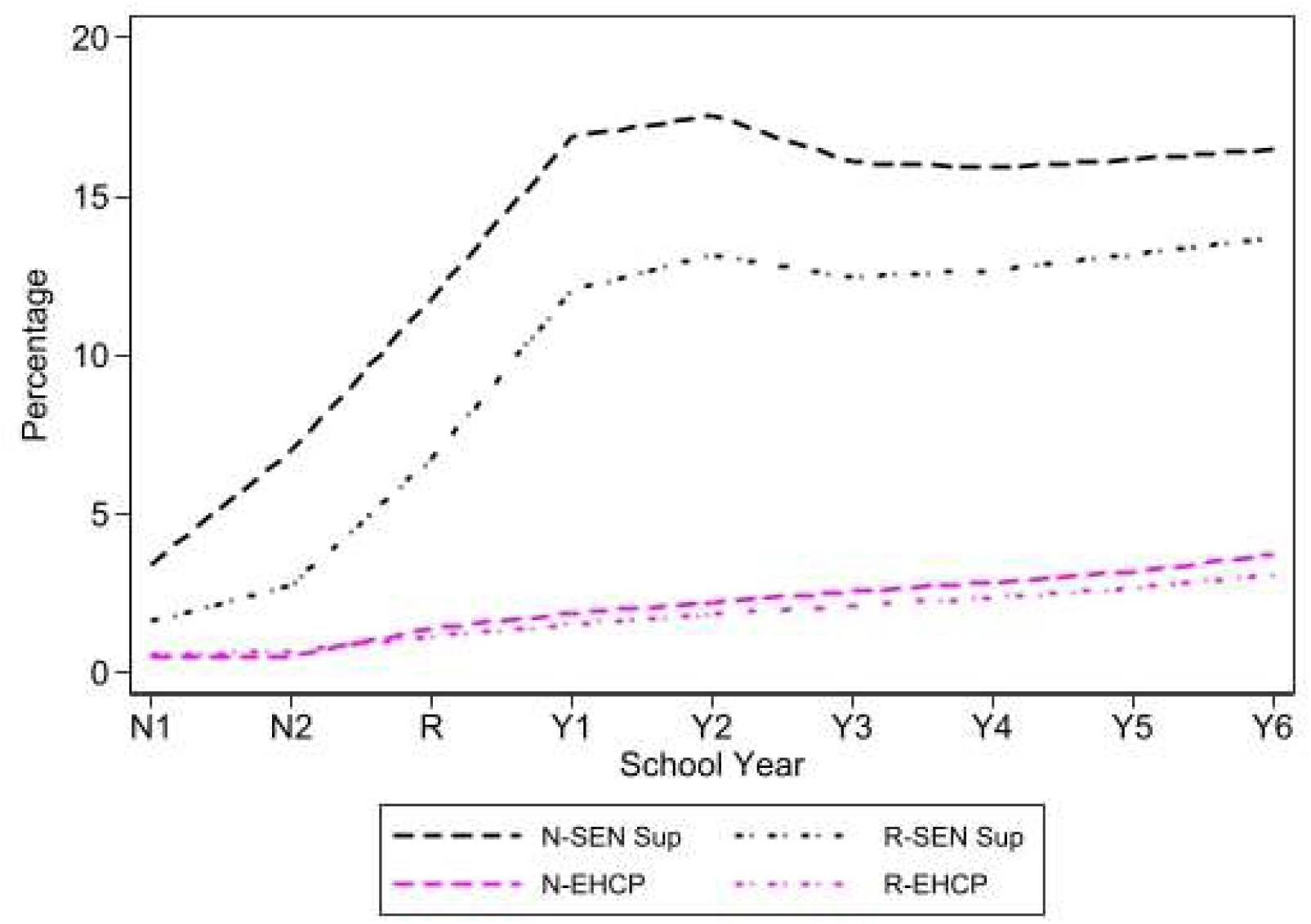
Cumulative percentage of children assigned SEN Support or EHCP provision during state-funded hours in Nursery or state primary school according to stage at entry to state education (Reception to Year 6);(Cohort 6, children born in 2006/07 or 2007/08) *Footnote: The percentage of children in each academic year who were assigned SEN Support or EHCP are shown by stage at entry into state education. N-SEN Sup and N-EHCP refer to children entering state education in Nursery and R-SEN Sup and R-EHCP refer to those entering in Reception class. The small proportion (0.4%) entering in Year 1 is not shown. As SEND provision during state-funded hours in a private or voluntary nursery is included, children entering state education in Reception can have SEND provision earlier (Appendix 4, paper 15).*[45]

In analyses restricted to high-need children with cerebral palsy (Cohort 8), we found no differences in the probability of an EHCP according to free school meal eligibility. However, we also found delayed EHCP provision for those living in the poorest neighbourhoods (Appendix 4, paper 12).[47]

### 2c. Does where the child lives and goes to school influence SEND provision?

The type of school governance has changed over time with a shift to more autonomous academy schools since 2007.[48] The 1 million children in Cohort 6 entered Year 1 in 2011/12 or 2012/13. At this point most children attended a community school (58.0%) with the rest attending voluntary aided (17.1%), voluntary controlled (9.2%), foundation (5.7%), academy converter (7.0%) or academy sponsor-led (2.6%).[45] A group comprising free schools and some special schools accounted for a further 0.3% of pupils but are excluded from further analyses (see Appendix 1).

We found substantial variation in the probability of SEN Support or EHCP provision in Year 1 across these different types of school governance.[45] Voluntary aided and voluntary controlled schools – typically religious schools – had the lowest rates of SEN Support or EHCP provision and academy sponsor-led and community schools had the highest rates (Figure 7). Variation diminished after equalising populations according to child-level differences in health characteristics, sociodemographic, stage at entry, and expected levels for the EYFSP assessment at age 5, but voluntary and academy sponsor-led schools had a lower probability of EHCP provision than community schools (Appendix 4, paper 15).[45]

**Figure 7.**
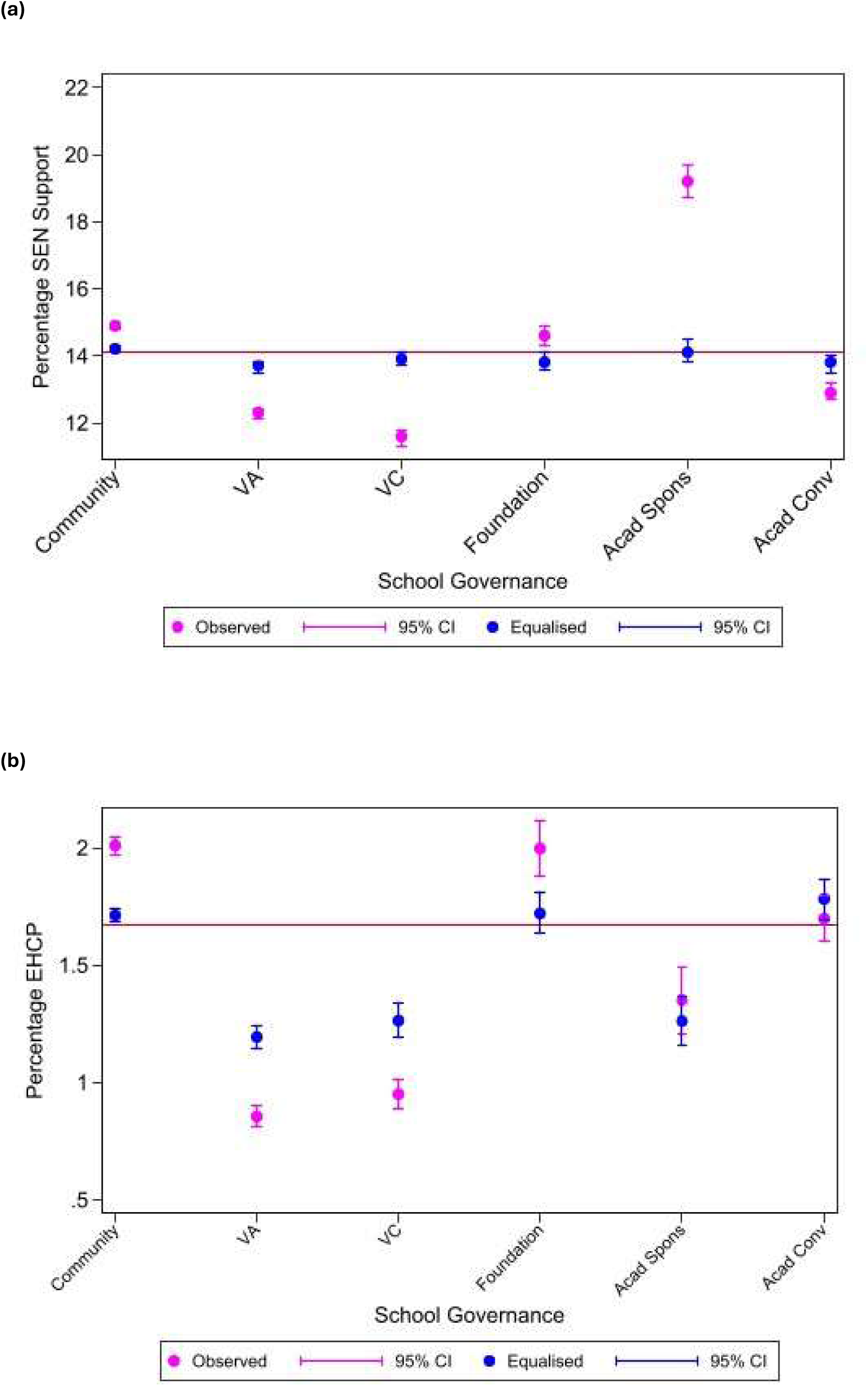
Observed and equalised probability of (a) SEN Support and (b) EHCP provision by school governance. (Cohort 6, born 2006/07 and 2007/08)

In separate analyses using cohort 7 (born 2003/4 to 2013/14), we found minimal variation in SEN Support compared with no SEND provision and EHCP compared with SEN Support across local authorities (Appendix 3, cohort 7).[49] After adjusting for child-level variables, only a small proportion of the variation was attributable to differences between LAs: it ranged from 2.0% for SEN Support (vs none) to 5.8 percent for EHCPs (vs SEN Support) across gestational age subgroups of cohort 7.[49] Including LA income deprivation in the model reduced the variance in EHCP provision by up to 24 percent (Appendix 4, paper 13).[49]

### 2d. Was SEND provision affected by the 2014 education reforms?

SEND provision declined steadily between 2009/10 and 2018/19. Figure 8 shows the prevalence of any SEND provision in Year 1 falling from 21.1 percent to 15.0 percent (Appendix 4, paper 9).[38] This declining trend was evident among children with major congenital anomalies as well as those without. Notably, there was no step change following the 2014 SEND reforms, thereby limiting the opportunity to use the occurrence of legislative change as an instrumental variable in a natural experiment design to evaluate the impact of SEND provision.[38]

**Figure 8.**
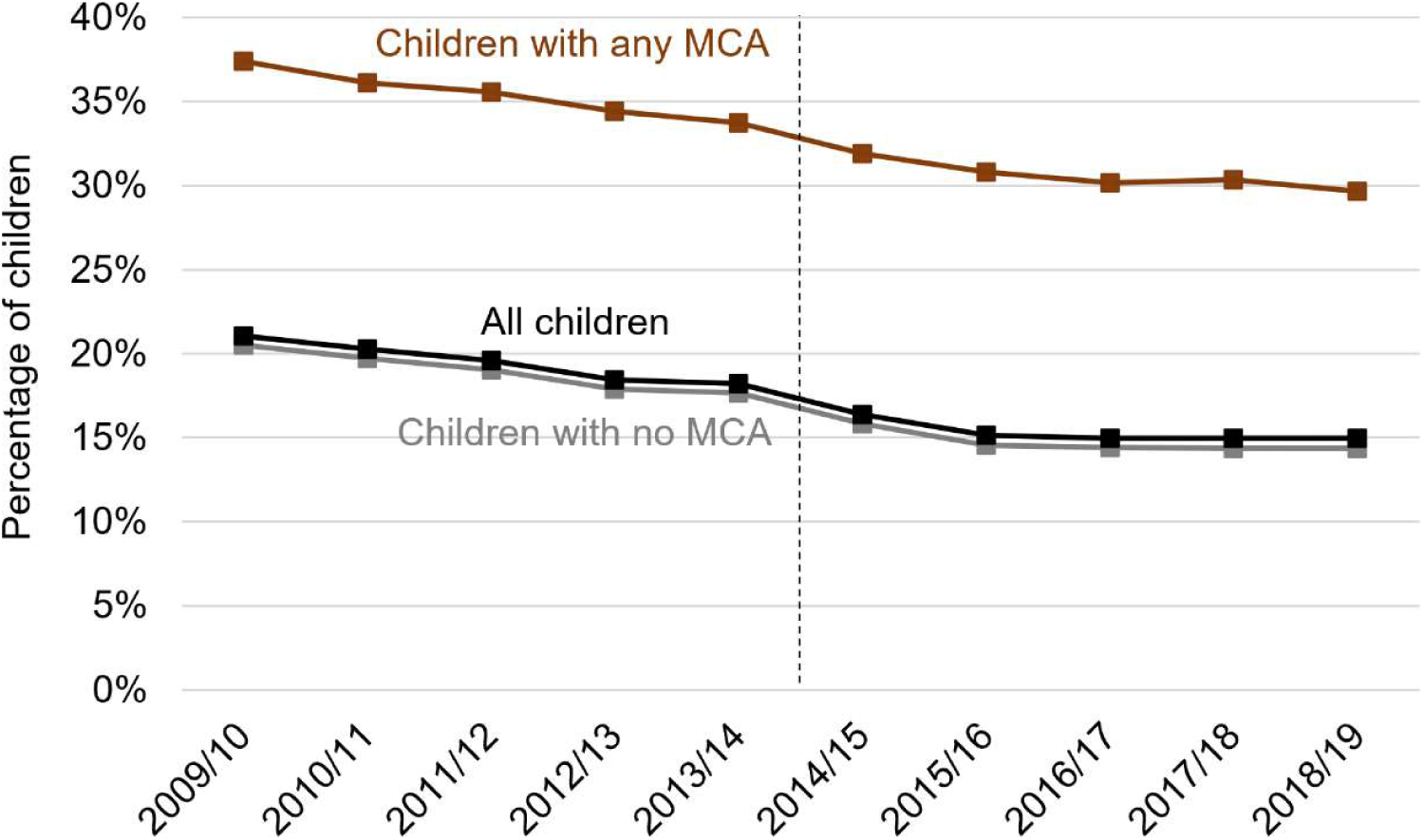
Prevalence of any SEND provision recorded in Year 1, by academic year and major congenital anomaly (MCA) status.

#### Question 2: Key Findings

##### Who is more likely to receive SEND provision?

- Children with higher-need health conditions, boys, summer-born children (youngest in class), and those from disadvantaged backgrounds were more likely to receive SEN Support or an EHCP.
- Children with neurodisability had the highest rates of SEND provision relative to children with congenital anomalies or those born preterm but provision varied widely by phenotype subgroup.
- Measures of chronic health conditions and gestational age at birth recorded in hospital records were not an adequate proxy of need for SEND provision. Hospital data alone does not adequately capture underlying need for SEND. Children with chronic health conditions made up 30.5% of children assigned any SEND in Year 1 and 63.9% of those with an EHCP.

##### When is SEND provision assigned?

- The probability of being assigned SEN Support or an EHCP increased only gradually from Nursery onwards, peaking in Year 2 for SEN Support and in Year 6 for EHCP
- Children who started in Nursery were more likely to receive SEN Support or an EHCP throughout primary school than those starting in Reception.
- Among children with cerebral palsy, 95% received SEN Support and 72% had an EHCP by the end of primary school.

##### Does where the child lives and goes to school influence SEND provision?

- Type of school governance was associated with SEND provision.
- Voluntary and academy sponsor-led schools had a lower probability of EHCP provision than community schools after accounting for pupil characteristics.
- Variation across LAs was minimal and mostly explained by child-level characteristics and LA area-level deprivation.

##### Was SEND provision afected by the 2014 SEND reforms?

- SEND provision in Year 1 declined gradually from 2009/10 to 2018/19 but there was no appreciable step change when SEND reforms were introduced.

### Question 3: What is the impact of SEND provision on health and education outcomes?

We found insufficient variation in SEND provision (or subcategories of support or EHCP) over time, across schools, local authorities, or among children (e.g. by month of birth), to use any of these features as valid instruments to deal with unmeasured confounding. Instead, we employed causal inference methods within a TTE framework to estimate the causal impact of SEND provision in two phenotypic cohorts: children with cleft lip with palate (cohort 9), and children with cerebral palsy (cohort 10). We estimated the impact of SEN Support versus none for both phenotypes, and EHCP versus SEN Support in Year 1 (for cerebral palsy) on unplanned admissions, unauthorised absences and key stage test scores through primary school (Table 1; Appendix 4, papers 16,18).[53][54]

**Table 1.** Summary of findings of causal analyses to estimate the impact of SEND provision on health and education outcomes (cohorts 9, 10; Appendix 4, papers 16,18).

| Phenotype and compared interventions in Year 1 | Unplanned admissions Year 2-Year 6 | Unauthorised absences Year 2-Year 6 | KS1 Year 2 | KS2 Year 6 |
| --- | --- | --- | --- | --- |
| **Cleft lip and/or palate**<br>(Appendix 3, cohort 10; Appendix 4, Paper 16) |  |  |  |  |
| SEN Supp vs No Support in Year 1 (Ref) | No difference | Better in SEN Supp | Worse in SEN Supp | Worse in SEN Supp |
| **Cerebral palsy**<br>(Appendix 3, cohort 9; Appendix 4, Paper 8) |  |  |  |  |
| SEN Supp vs No Support in Year 1 (Ref) | Worse in SEN Supp | Better in SEN Supp | Worse in SEN Supp | No difference |
| EHCP vs SEN Support in Year 1 (Ref) | Worse in EHCP | Better in EHCP | No difference | Worse in EHCP |

We found limited evidence that early SEN Support vs none improved rates of unplanned hospital admissions or educational attainment in either the cerebral palsy or the cleft lip and/or palate phenotype subgroups. Findings were similar for EHCP vs SEN Support for children with cerebral palsy.[53] [54] However, there was evidence that SEN Support compared to no support reduced the rate of unauthorised absences for both phenotype groups. Among children with cerebral palsy, both comparisons (SEN Support vs none, and EHCP vs SEN Support) revealed reduced unauthorised absences, although there were no differences in rates in total absences. [54]

The findings of improved and worse outcomes in Table 1 should be interpreted with caution given the likely impact of residual confounding arising from the coarseness of the measures available in ECHILD. Furthermore, our choice of outcomes was limited by the accuracy and relevance of data available. For example, absences may be less likely recorded to be recorded as unauthorised for children with an EHCP. Also, other outcomes relevant to learning or socio-emotional skills may not be captured in ECHILD.[53] [54]

#### Question 3: Key Findings

- We found insufficient variation in SEND provision across time, place, and child subgroups to use instrumental variables to account for unmeasured confounding in natural experiment designs.
- Using the TTE framework and propensity score-based and g-methods, we were able to examine and justify the key assumptions (for positivity and conditional exchangeability) only for narrowly defined phenotypic subgroups (cleft lip with palate and cerebral palsy), and for certain comparisons (recorded provision in year 1, comparing SEN Support vs none, on outcomes from Year 2 to the end of Year 6).
- We found no evidence that SEND provision in Year 1 reduced hospitalisation rates or improved attainment.
- SEN Support did appear to reduce the rates of unauthorised absences in comparisons to no support for both phenotypes, with reduction also found for EHCP vs SEN Support in the cerebral palsy phenotype.
- Our results should be interpreted cautiously as unmeasured confounding might affect both the null results and the reported benefits of SEND provision (as not all indicators of need were measured).

### Question 4: What do service experiences and policies tell us about SEND processes?

Ten mixed-methods studies informed the design and interpretation of the ECHILD analyses. Findings are summarised below for initial identification (4a), assessment and planning (4b), provision (4c), outcomes (4d), local service capability (4e), and national policy context (4f). Appendix 5 summarises and maps methods and findings from each study onto questions 4(a to f). Further details are in papers in Appendix 4, papers 21-30).

### Initial identification (4a)

In qualitative studies, professionals reported increasing numbers and complexity of the need for SEND provision, linked to worsening poverty and in some settings, better identification by experienced Special Educational Needs Coordinators (SENCOs).[66] Parent-carers reported that quiet children, and autistic girls were frequently under-identified.[67] Online surveys found that 25% of children had been identified in pre-school and 47% by Key Stage 1. Parents reported negative experiences of identification, inadequate professional training, long waits, and limited specialist availability.[68] [69] While most professionals felt confident in identification, they mostly agreed that interagency communication was weak.[70] Document reviews revealed variation by multi-academy trust (MAT), SENCO level of experience, and LA. Many Local Offer (LO) websites lacked clear, accessible criteria and process information.[71][72][73][74]

### Assessment and planning (4b)

EHCP processes were described in qualitative studies involving professionals as inefficient, due to poor interagency working, information sharing systems, and a lack of shared understanding of SEND provision and processes. Parents reported that high intervention thresholds and referral bounce-back harmed relations with families and left children unsupported, sometimes for years.[66][67][75] Surveys found fewer than one-quarter of parents reported adequate information at assessment; only half of children felt involved in decisions.[68][69] Less than half of professionals reported that parents unable to advocate were adequately supported through the SEND process.[70] Document reviews highlighted weakened EHCP quality due to inconsistent application of Children and Families Act (2014) legislation, varying criteria for some SEND types, funding-driven allocation, poor co-production and involvement of children, poorly defined outcomes, and incomplete LO website eligibility criteria. Poor quality EHCP processes and provision were common reasons for LAs failing inspections.[71][72][73][74]

### SEND provision (4c)

In the qualitative studies, we heard frequent reports of delayed and inappropriate SEND provision, harming children’s education and mental health. Tailored support depended on good SENCO training. Parent-carers acted as primary advocates but were excluded from LA decision-making panels. Appeals improved provision but were slow and costly; annual reviews were uncommon.[66][67][75] Surveys found 75% of children wanted earlier support; only half reported inclusion similar to peers. Only one-third of parents said provision matched plans, and 41% said plans matched needs.[68][69] Only 45% of professionals rated SEND provision as positive: their level of confidence was mixed in being able to deliver SEND provision or having the capacity to support families.[70] Document reviews indicated that LAs were frequently under-resourced for needs-led delivery.[71][72][73][74]

### Outcomes (4d)

Young people commented in qualitative studies that timely, tailored provision improved academic engagement, anxiety management, and independence. Challenging unmet needs took a toll on parent-carer mental health and finances. Professionals sometimes set unduly low aspirations. Transitions required careful management, and reliance on parental advocacy reinforced inequities.[66][67][75] In surveys, only half of parents reported that EHCPs included future goals; only one-third found these appropriately ambitious.[68][69] Document reviews associated poor provision with dissatisfaction, complaints, and disproportionate exclusions.[71][72][73][74]

### Local service capability (4e)

Qualitative studies indicated capacity constraints due to workforce shortages, insufficiently trained staff, complex caseloads, funding deficits, inconsistent eligibility thresholds, and limited specialist availability. High-quality information systems improved interagency working but were rare.[66][67][75] Surveys found low awareness of what SEND provision was available locally (the LO), high parent-carer dissatisfaction with LA assessment processes reflected by 38% of parents reporting that they paid for private assessments.[68][69] Document reviews confirmed variation in local capacity, leadership, funding, EHCP rates, consistency of processes and interagency working. Nationally, there are increasing complaints to the Local Government and Social Care Ombudsman (LGSCO) in an increasing number of LAs.[71][72][73][74]

### National policy context (4f)

Qualitative studies found that a narrow curriculum, exam-focused assessment, academisation, rigid attendance policies, and specialist shortages undermined inclusion and set some children up for failure. Educators were increasingly absorbed in mental health roles, reducing capacity for prevention.[66][67][75] Professionals reported that insufficient funding and training reduced service functioning and capacity.[68][70] Document reviews found that professionals wanted nationally standardised processes and guidelines to improve quality. National policies on academisation reduced oversight and accountability for poor school practices on SEND provision. Some reports commented that the SEND code of practice led to under-recording of need, reduced the supply of specialist teachers and did not ensure LA compliance with legal requirements on the LO.[71][72].

## Discussion

### Who is assigned SEND provision, when and where? What do service experiences and policies tell us about the SEND process?

Nearly one-third (30%) of children who entered primary school were assigned SEND provision by age 11 in 2014-19. SEND provision was strongly linked to health characteristics such as having a chronic health condition, including neurodisability or a congenital anomaly, or being born too early. However, these characteristics alone were not sensitive indicators of SEND provision. Other factors, such as being male, facing social disadvantage, and a low developmental score at age 5, were also important.

When children were assigned SEND provision was complex. Children with social disadvantage or disability, which is linked to eligibility for state-funded nursery at age 2-3,[6] had higher rates of SEN Support and EHCP provision throughout primary school than their peers. Despite this increased provision, many did not start SEND provision until Year 1 or 2, possibly due to delayed recognition, lack of school capacity, or changing need as education became more complex. However, in analyses of children with cerebral palsy, we found that those entering state education in Nursery and those living in the 20% most income-deprived neighbourhoods were less likely to be assigned EHCPs than their peers.[47]

Which type of school children attended was associated with their chances of SEND provision. Children attending voluntary, religious schools and academy sponsor-led schools were less likely to be assigned an EHCP compared with community, foundation and academy converter schools.[45]

Findings from ten mixed methods studies highlighted variability in services. Some children and families reported meaningful benefits from provision, however, interactions with services, delays and inadequately tailored interventions frequently included negative experiences, some experiencing long term harm. Qualitative studies and surveys revealed unmet need and delays in identification of need, assessment, planning and provision, that particularly affected disadvantaged children who did not have parent-carers who could advocate for them. Parent-carer, child and professional perspectives and document reviews reflected services that lacked capacity, training and specialist expertise. These findings also highlighted a system that interacted poorly with other weakened systems (such as CAMHS) and lacked national guidance and accountability. These problems were compounded by underfunding, academisation, and a narrow curriculum that was perceived to undermine inclusion of children’s diverse abilities.

### What is the impact of SEND provision?

We found weak evidence that SEND provision in Year 1 (age 5/6) reduced rates of unauthorised absences, but no evidence for reductions in hospital admissions or improved attainment during primary school. These equivocal results are consistent with 49 natural experiment studies worldwide that evaluated SEND provision as implemented in routine practice (see introduction).

These quantitative findings were complemented by qualitative findings in the HOPE study from parent-carers and children using SEND services and professionals. Their reports indicated that timely, tailored provision improved academic engagement, anxiety, and independence for children. However, the stresses of the SEND process, including inappropriate or inadequate provision, were experienced as harmful for both children and their families.

### Strengths and limitations

Strengths of the HOPE study included exploration of the evaluability of SEND provision using different approaches to causal analyses using rich, longitudinal health and education data for all children in state-funded primary education in England captured in the ECHILD database. Use of mixed methods and stakeholder engagement, alongside the quantitative analyses, were critical for understanding variation in the adequacy of SEND processes and how many elements were not measured in administrative data. Limitations centred on a data science problem: inadequate measures of the need for SEND and other confounders, limited information on the nature of the intervention, and insensitive or insufficiently relevant outcomes that might not be measurably changed by SEND provision. A further limitation was that we found no instrumental variables to use in natural experiment designs that address unmeasured confounding, despite major changes in national SEND policy and types of school over the past two decades. These limitations undermined the evaluability of the impact of SEND provision on health and education using quantitative analyses of ECHILD.

### Challenges and opportunities in using administrative data

Our findings reflect challenges that are relevant to other jurisdictions that use administrative data to evaluate SEND provision. One lesson from the HOPE study is that health phenotypes as recorded in hospital data do not adequately proxy need for SEND provision. Linked hospital and education data lack direct measures of function, disease, severity or complexity of need, and social and behavioural factors linked to high need and worse outcomes.

Secondly, SEND provision recorded in ECHILD reflects assignment to the SEND process but not what activities or support were received, at what intensity and when. Young people and parent-carers reported frequent delays in receiving support, watered-down activities or activities the child did not tolerate, due to lack of capacity, skills or ability of staff to adapt activities to the child.

Third, the outcomes measurable in ECHILD - hospital admissions, attainment test results and school absences - are relatively insensitive to positive changes reported in response to SEND provision by parent-carers and young people. They mentioned feeling happier, improved sense of self, feeling less anxious or distressed, improved behaviour and mental health, and being able to attend school, participate, learn and socialise. Impacts on parent wellbeing were also mentioned, while reduced hospital contacts for the child were not.

Data linkage to relevant information from administrative, cohort, survey, trial or audit data on need for, and content of, SEND provision, and on relevant outcomes, could mitigate these three challenges.

Fourth, the SEND process may have been harmful for some children and families. Our study spanned a period of national austerity, with cuts to education and related services, rising child poverty and need for SEND provision. These changes occurred in parallel to policy changes that incentivised academic attainment and reduced funding and accountability for progress among children with additional needs. Parents and practitioners reported erosion in the capacity or skills to deliver SEND provision of adequate quality. These problems created distrust between parents and school staff and undermined partnership working between staff, child and parent-carer.

Fifth, notwithstanding these limitations, the HOPE study illustrates the opportunities to inform policy makers, teachers, clinicians and families about children’s trajectories through health and education and into adulthood. Descriptive analyses while being essential to design causal analyses, also provided important information on expected outcomes and variation and inequalities in SEND provision.

### Implications for policy

Achieving effective and equitable SEND provision is a policy challenge in many countries. SEND provision is an expensive and important service - £11 billion per year in England - with the potential to change the life opportunities of one-third of all children.[31][76] More effective and efficient services could improve experiences and outcomes for children and their families, limit escalating costs, and yield long-term benefits across public services as better supported, more independent and educated children with additional needs transition to adulthood. Broader policy changes are also needed beyond SEND provision, including broadening the academic curriculum to value and measure other forms of progress, including social and emotional skills. Beyond schools, better integration is needed between health and education services for children with additional health and social needs, and more support for parent-carers to reduce the toll on families.

Further reforms to SEND in the UK and elsewhere, [77] [78] need to be based on, and continue to generate, evidence of what works. Development of high quality linked data research infrastructure across education and other public services should be at the forefront of government’s research areas of interest.[77]

### Implications for research

Despite the hundreds of RCTs of targeted SEND interventions and, with this study, 50 natural experiment studies on the impact of SEND provision, uncertainty remains about what works for whom, in what contexts and how. Achieving effective SEND provision requires randomised trials of alternative approaches to SEND provision in situations where uncertainty exists and comparators are acceptable. For example, the HOPE study found that SEND is provided to few children in the general population in the preschool years (Nursery and Reception class).

Although earlier implementation makes intuitive sense, and is provided for some disabilities such as visual impairment, there is limited evidence for broader needs from contemporary nursery-based trials.[79][80] A randomised, staggered trial could be used to implement planned policy change in England and, at the same time, generate robust evidence on early versus deferred SEND provision in the early years. [81]

Policy makers also need evidence on what good SEND provision and alternative practice options look like to develop meaningful evaluations of impact. Our mixed methods studies revealed a SEND service that few users experienced as being able to meet needs due to lack of funding, staff, skills, interagency support and trust from families.[31] Comparison of such a weakened system of SEND provision against education as usual is unlikely to advance policy.

More and better research is needed to guide development of effective and equitable SEND provision globally. International collaboration could accelerate delivery of research for example, sharing knowledge on administrative data linked across education, health and other sectors to enable parallel studies across diverse needs and contexts relevant to SEND provision. Surveys, cohort studies, and RCTs of SEND provision, or targeted components, in education or healthcare, could be conducted and linked into administrative data. Such linkages would minimise costs of data collection, and if conducted across jurisdictions, could improve comparability and give insights about differing impacts according to context. Sharing of mixed methods approaches would improve understanding of services across jurisdictions.

International research collaboration is needed to build on the findings from the HOPE study through wider integration of natural experiment, randomised and mixed methods studies to improve the effectiveness of SEND provision. However, the challenges facing the HOPE study, are echoed by other population-based, service interventions, such as early home visiting for first-time, teenage mothers and perinatal mental health hubs.[82] [83][85] The recent framework on use of natural experimental designs,[21] needs to be extended to consider how policy makers, funders, and researchers can introduce and document changes in policy and practice and efficiently enhance administrative data to enable robust evaluation of education, health and social interventions in populations.

## Data Availability

The ECHILD database is made available for free for approved research based in the UK, via the ONS Secure Research Service. Enquiries to access the ECHILD database can be made by emailing. Researchers will need to be approved and submit a successful application to the ECHILD Data Access Committee and ONS Research Accreditation Panel to access the data, with strict statistical disclosure controls of all outputs of analyses.

https://www.echild.ac.uk/data-access

## Acknowledgements

**Members of the HOPE study team include**: Ruth Gilbert (PI), Katie Harron, Bianca L De Stavola, Lorraine Dearden, Tamsin Ford (senior work package leads), Kate Lewis, Vincent Nguyen, Ania Zylbersztejn, Jennifer Saxton, Jacob Matthews, William Farr (leading roles in the 4 work packages), Ayana Cant, Laura Gimeno, Isaac Winterburn, Ariadna Albajara Saenz, Andrea Aparicio Castro, Julia Shumway, Lucy Karwatowska, Ananya Khera, Nicolas Libuy, Louise Macaulay (contributing researchers), Matthew Lilliman (programme manager), Kate Boddy (public engagement coordinator), Stuart Logan, Jugnoo Rahi, Kristine Black-Hawkins, Johnny Downs, Kristine Black Hawkins (co-investigators).

We are grateful to the HOPE study Advisory Group: Chris Bonell (chair, professor of health and sociology, London School of Hygiene and Tropical Medicine), Kate Evans-Jones, Julia Ogden (public members), Jo Hutchison (Director of SEND, Education Policy Institute), Karen Horridge (consultant community paediatrician, Sunderland). Also thanks to Jo Van Herwegen (UCL IoE) for discussions and comments.

We gratefully acknowledge all children and families whose de-identified data were used in this research. We thank Ruth Blackburn, Milagros Ruiz, Matthew Jay, Antony Stone and Farzan Ramzan for ECHILD Database support.

The ECHILD Database uses data from the Department for Education (DfE). The DfE does not accept responsibility for any inferences or conclusions derived by the authors. This work contains statistical data from ONS which is Crown Copyright. The use of the ONS statistical data in this work does not imply the endorsement of the ONS in relation to the interpretation or analysis of the statistical data. This work uses research datasets which may not exactly reproduce National Statistics aggregates. Evidence from this research contributes to the NIHR Children and Families Policy Research Unit but was not commissioned by the NIHR Policy Research Programme.

## Funding

This project is funded by the National Institute for Health Research (NIHR) under its ‘Programme Grants for Applied Research Programme’ (Grant Reference Number NIHR202025, The HOPE Study). The views expressed are those of the authors and not necessarily those of the NIHR or the Department of Health and Social Care. ECHILD is supported by ADR UK (Administrative Data Research UK), an Economic and Social Research Council (part of UK Research and Innovation) programme (ES/V000977/1, ES/X003663/1, ES/X000427/1).

## Statement of Conflicts of Interest

Tamsin Ford’s research group receives funding for research methods consultation from Place2Be, a third sector organisation that provides mental health and training to UK schools.

The other authors report no conflicts.

## Ethics statement

Existing research ethics approval has been granted for analyses of the ECHILD database version 1 for the purposes set out in the HOPE study (20/EE/0180). Permissions to use linked, de-identified data from HES and the NPD were granted by NHS England (DARS-NIC-381972-Q5F0V-v0.5) and the Department for Education (DR200604.02B). Patient consent was not required to use the de-identified data in this study.

## Notes

### Competing Interest Statement

TF declared consulting fees from PLACE2Be
Funding declared from NIHR Programme Grant

### Clinical Protocols

https://pubmed.ncbi.nlm.nih.gov/37918923/

### Funding Statement

The study was funded by the National Institute for Health Research (NIHR) under its Programme Grants for Applied Research Programme (Grant Reference Number NIHR202025, The HOPE Study).

### Author Declarations

Ethical approval for the ECHILD project was granted by the National Research Ethics Service (17/LO/1494), NHS Health Research Authority Research Ethics Committee (20/EE/0180 and 21/SW/0159). Separate ethical approval was not required to use de-identified data for the analyses presented in this paper for the HOPE study (20/EE/0180). Permissions to use linked, de-identified data from HES and the NPD were granted by NHS England (DARS-NIC-381972-Q5F0V-v0.5) and the Department for Education (DR200604.02B). Patient consent was not required to use the de-identified data in this study.

