## Supplementary materials for "Evaluation of special educational needs and disability provision in English primary schools using administrative health and education data in the ECHILD database"

### Appendix

### Appendix 1: Glossary and policy context.

Table S1: Glossary of common terms and variables used in the HOPE study.

| | |
|---|---|
| **1. Phenotypes** | |
| **Neurodisability** | Chronic conditions involving impairment of the brain and/or neuromuscular system that result in functional limitations.[1] The list of conditions was informed by a scoping review of existing literature and refined through iterative clinical input. The full list conditions is available in the ECHILD phenotyping library.[2] |
| **Congenital anomalies** | Structural or functional birth defects that develop prenatally and serve as reliable early indicators of varying levels of need for SEND provision.[3] Subgroups were defined using the EUROCAT guide (version 1.5); we focused on a selection of major congenital anomalies categorised into 12 body system groups and 25 specific subgroups.[3] |
| **Chronic health conditions (CHC)** | Any health problem recorded in hospital admission data likely to require follow-up for more than 1year, where follow-up could be repeated hospital admission, specialist follow-up through outpatient department visits, medication or use of support services.[4] We identified children with CHCs using the Hardelid code list and the full list of conditions is available in the ECHILD phenotyping library.[2][4] |
| **Gestational age** | We stratified the whole population of children by week of gestational age at birth to assess the gradient in health and education outcomes across different levels of underlying need. Due to small sample sizes, individuals born before 24 weeks or after 42 weeks of gestation were grouped together, as counts below 10 are suppressed or treated as confidential to protect individual privacy. |
| **2. School governance and types of primary schools** | |
| **Inclusive education** | An approach to teaching that ensures all learners, regardless of ability, background, or need, have equal access to high-quality education, in line with UK laws such as the Equality Act 2010 and the SEND Code of Practice (2015).[5] The core principles are valuing diversity as a strength, removing barriers to participation, fostering social cohesion, addressing attainment gaps, and preparing students with the skills and attitudes needed to thrive in a diverse society. [6] |
| **Special school** | Special primary schools in England serve pupils with a broad spectrum of special educational needs and disabilities (SEND; see section 5 in this glossary), particularly those with moderate to severe or complex disabilities. These schools provide tailored support and specialised learning environments designed to meet the needs of children who require more intensive or individualised interventions than those typically available in mainstream educational settings.[7] Almost all children in a |

<table>
<tr><td></td><td>special school have an education, health and care plan (EHCP).[8] Special schools may be maintained by local authorities or operate as academies, including both sponsor-led and converter academies. They may be either faith-based or secular, and in exceptional cases, may operate as independent (private) schools.[9]<br><br>Alternative provision (AP) refers to education arranged by local authorities for children of compulsory school age who cannot attend mainstream or special schools, often due to behavioural, medical, or other specific needs. The most common form of AP is the Pupil Referral Unit (PRU), which supports pupils who are excluded or otherwise unable to attend a school setting. Local authorities have a statutory duty to ensure suitable AP is provided in these circumstances. [10]</td></tr>
<tr><td colspan="2">Local Authority Maintained Schools<br>Local authority maintained schools are fully owned and operated by local authorities. They follow the national curriculum, are closely connected to their local communities, and may offer additional services such as childcare. Maintained schools can be named in an EHCP (previously termed Statement of SEN), following consultation with the local authority, and must comply with government regulations on admissions, exclusions, and SEND provision.[11]</td></tr>
<tr><td>Community school</td><td>A local authority-maintained, secular school. The local authority manages admissions, staffing, and owns the buildings.[11]<br>The proportion of community primary schools has reduced over time as Foundation and Academy Trust schools have increased from 2007 onwards.[12]</td></tr>
<tr><td>Voluntary Aided school</td><td>These schools are typically faith-based, though admissions are open to all applicants. They are jointly funded by the local authority and a supporting religious body, such as the Roman Catholic Church. The governing body employs staff, sets admissions policies, and contributes to building and maintenance costs. School premises are usually owned by a charitable foundation.[11]</td></tr>
<tr><td>Voluntary Controlled school</td><td>Similar to voluntary aided schools, these are mostly faith-based but are fully funded by the local authority. While the local authority is responsible for admissions and employs school staff, it consults with the supporting body when setting the admissions policy. The land and buildings are typically owned by a charitable foundation.[11]</td></tr>
<tr><td>Foundation school</td><td>Funded by the local authority but are managed by their governing body, which acts as the admission authority. The governing body is responsible for employing staff and overseeing admissions. The land and buildings are owned either by the governing body or a charitable foundation.[11]</td></tr>
<tr><td colspan="2">Single Academy Trust (SAT) / Multi-Academy Trust (MAT)<br>A SAT is a standalone academy (sponsor-led or converter) with its own board of trustees, independent of local authority control and funded by the Department for Education. A MAT is a group of academies governed by a single board of trustees, sharing policies, leadership, and resources; MATs may include primary, secondary, and special schools. The HOPE study does not distinguish between schools under SATs or MATs.[13] [14]</td></tr>
</table>

| | |
|---|---|
| **Academy (Sponsor-led)** | State-funded school set up to replace underperforming schools. Overseen by an external sponsor (e.g., trust, charity). Operates independently from local authorities with freedom over curriculum, staffing, and budget.[9] [14] |
| **Academy (Converter)** | A higher-performing school that voluntarily converts to academy status. Funded directly by the Department for Education and has autonomy over operations and curriculum. [9] [14] |
| **3. Methodology** | |
| **3.1 Quantitative methods** | |
| **Natural experimental evaluation** | Natural experiments are defined as events outside the control of researchers that divide populations into exposed and unexposed groups.[15] They pose a valuable opportunity to evaluate population health, health systems, and other interventions, including those that are, for practical or ethical reasons, not suitable for investigation using randomised controlled trials. A natural experimental evaluation uses differences in exposure generated by an event or process that arises independently of the outcome to identify, measure or understand the effects of that exposure.[15][16] |
| **Instrumental variables** | A variable that is associated with exposure to an intervention but does not have a direct association with the outcome of interest. An instrumental variable can be used to estimate the effect of exposure on an outcome if: (1) it is associated with the outcome but (2) only through its association with the exposure (the 'exclusion restriction') and (3) it is unrelated to any other factors that cause the outcome.[15] |
| **Intraclass correlation coefficient (ICC)** | Measures the consistency or agreement of measurements within grouped data. It quantifies how much of the total variation is due to differences between groups versus within groups, indicating how similar members of the same group are compared to those in different groups.[17] |
| **Target trial emulation (TTE)** | A framework that applies the study design principles of randomised trials to observational studies that aim to estimate the causal effect of an intervention. This approach aims to reduce bias and strengthen causal validity in observational studies.[18] |
| **3.2 Qualitative methods** | |
| **Focus group discussions** | A focus group discussion (FGD) is a qualitative method where a moderator guides a selected group to discuss a topic in-depth. FGDs capture participants' attitudes, knowledge, and experiences through group interaction, revealing both shared understandings and diverse perspectives that might be missed in individual interviews.[19] |
| **4. Datasets** | |

| | |
|---|---|
| **Education and Child Health Insights from Linked Data (ECHILD)** | A pseudonymised database which links administrative data from the National Pupil Database (NPD) and Hospital Episode Statistics (HES).[20] The linkage rate between NPD and HES is high and improved with time (94-98%).[20] [21] The version of ECHILD used in the HOPE study analyses contains data on approximately 14.7 million children and young people aged 0-24 in England who were born in NHS-funded hospitals between 1st September 1995 and 31st August 2020. [21] |
| **4.1 Health data** | |
| **Hospital Episode Statistics (HES)** | A national database that includes information on all National Health Service (NHS) acute hospital care and mortality data. Nearly all children born in England are born in NHS hospitals (97%), but HES excludes births in private hospitals or at home. Children can be followed from their birth admission through all subsequent NHS hospital contacts.[21] |
| **HES Admitted Patient Care (HES APC)** | Data that is collected on all admissions to NHS hospitals in England and is the main HES data module used in HOPE study analyses. A hospital admission includes any secondary care-based activity that requires a hospital bed, thus includes both emergency and planned admissions, day cases, births and associated deliveries. [22] |
| **International Classification of Diseases version 10 (ICD-10)** | A standardised medical classification system developed by the World Health Organization.[23] In NHS hospitals, clinical coders use ICD-10 codes to record patient diagnoses based on care records or discharge summaries.[22] In HES APC data, diagnostic codes follow the ICD-10 system, which enabled us to identify cohorts of children with specific phenotypes of interest (e.g. chronic health conditions, including neurodisability or congenital anomalies) in analyses.[21] |
| **Office of Population Censuses and Surveys Classification of Interventions and Procedures version 4 (OPCS-4)** | A UK-specific statistical classification system used to code clinical procedures and interventions carried out in NHS hospitals.[24] OPCS-4 codes are recorded in HES APC data and were used in HOPE to identify groups of children with a phenotype of interest. |
| **4.2 Education data** | |
| **National Pupil Database (NPD)** | A record-level administrative data resource curated by the UK government's Department for Education that is used for funding purposes, school performance tables, policy making, and research.[25] NPD contains child-level and school-level data on all pupils in state schools in England. The primary module is the census, which has information on pupil characteristics and school enrolment. Other modules include alternative provision, exam attainment, absence and exclusions.[25] |
| **Early Years Census** | The Early Years Census (EYC) is an annual data collection by the Department for Education (DfE) in England. It captures information on children under 5 receiving free state-funded early education hours in various settings, including school-based nurseries, state-funded nurseries not based in schools and private, voluntary, or third-sector nurseries delivering state-funded hours. Data is collected each January and includes |

| | |
|---|---|
| | information on children's age, gender, and ethnicity, as well as provider details.[26] |
| **5. School variables** | |
| **Early Years Childcare Providers** | Settings offering care and education for children aged 0–5, including nurseries, pre-schools, childminders, and school nursery classes. All 3- and 4-year-olds are entitled to 15 hours of free childcare (up to 30 hours for eligible families), which can be used at both state-funded and private providers. Private nurseries may charge for extras (e.g. meals, extended hours), even during funded hours, while state-funded nursery classes (in schools) typically do not.[27] [28]<br><br>From 2014, children in families receiving welfare benefits, or with disability living allowance or an EHCP, were entitled to 15 free hours free nursery provision for 38 weeks a year from the age of 2. |
| **Academic school year** | Runs from 1st September to 31st August in England. Determines school year group placement and eligibility for assessments and services.[29] |
| **Compulsory schooling** | Children must start full-time education in primary school on 31 December, 31 March or 31 August following their fifth birthday - whichever comes first.[30] |
| **Stage at start of education** | The stage at education (see Question 2b, paper 15, cohort 6) was defined as the year of starting state-funded education. This could be state-funded free hours in early years education, such as Nursery, or starting state-funded primary school (Reception class or Year 1). |
| **Early Years Foundation Stage Profile (EYFSP)** | A statutory assessment of children's development at the end of the Reception year (age 4/5). Teachers assess each child against 17 Early Learning Goals (ELGs) across 7 areas of learning (2012/13 onwards). For each ELG, children are judged as either meeting the expected level or 'emerging' (not yet meeting it). The Profile provides a reliable summary of development to support a smooth transition into Year 1.[31] |
| **Good Level of Development (GLD)** | A child is defined as having achieved the GLD in the EYFSP if they have reached the expected level of development in the prime areas of learning (communication and language, physical development, and personal, social and emotional development) and the specific areas of literacy and mathematics.[31] EYFSP analysed in different forms. i) In Question 2 (Appendix 4, paper 15) EYFSP was standardised to a mean score and then categorised to a) not recorded, b) below the mean; c) mean or above. ii) in Question 1, Appendix 4, paper 7, it was analysed as a standardised mean score and also as a dichotomised variable: a) achieving a Good Level of Development; b) not achieving or not being assessed. Iii) In question 1 Appendix 4 paper 5, dichotomised as a good level of development (as above in ii(b)). |
| **Key Stage 1 and 2 assessments (KS1, KS2)** | Key Stages are phases in the national curriculum in England, each covering specific school years with standardised subjects and assessments – also termed attainment. At the end of Year 2 |

| | |
|---|---|
| | (age 6–7), children take KS1 assessments in English (reading and writing), maths, and science. At the end of Year 6 (age 10–11), KS2 assessments cover English (reading, writing, grammar, punctuation and spelling), maths, and science.[32] KS1 assessments are marked locally in schools while the KS2 assessments are externally marked. [33] [34] Students who did not follow the National Curriculum (disapplied) or who were assessed according to alternative standards (P-scales), were considered as "not assessed" for our study. |
| **Authorised absences** | The number of school sessions (half-days) a child misses in an academic year with the school's permission. These absences are reported by schools and may be due to illness, medical or dental appointments, religious observance, study leave, agreed family holidays, or other approved reasons.[25][35] |
| **Health-related absences** | An authorised school absence recorded due to illness or medical/dental appointments, as classified in the NPD. Used as a health outcome measure in studies of children with neurodisability (Paper 18).[36] |
| **Unauthorised absences** | The number of school sessions (half-days) a child misses in an academic year without permission from the school. This includes unexplained or unjustified absences such as truancy, unapproved holidays during term time, lateness, and absences where no reason has been provided.[25][35] |
| **Persistent absence** | Refers to pupils who miss more than 10% of possible school sessions in an academic year, regardless of whether the absences are authorised or unauthorised.[25][35] |
| **Special Educational Needs (and Disabilities)**<br><br>*Changes in language throughout the HOPE study. | The Special Educational Needs and Disability Act 2001 and the SEN Code of Practice (2001) define children with special educational needs as those who have a learning difficulty requiring special educational provision.[37] This includes difficulties in learning as well as disabilities. The Children and Families Act (2014) and the updated SEND Code of Practice use a similar definition but explicitly adopt the term SEND to encompass both special educational needs and disabilities.[38][5] The SEND system is the formal framework used in England to identify children and young people who *'have a learning difficulty or disability which calls for special educational provision, has a significantly greater difficulty in learning than the majority of others of the same age, or has a disability which prevents or hinders him or her from making use of facilities of a kind generally provided for others of the same age'* (See page 15, SEND code of practice).[5] SEND provision refers to both types of support a child may receive under this system (SEN Support or EHCP).<br><br>*In the HOPE study, we initially used the term 'SEN provision' to remain consistent with NPD. However, we shifted to 'SEND provision' to align with current policy language and to ensure that disabled children, especially those in mainstream education, are not overlooked or excluded.[5] Not all children with disability will have SEN; for example children with myopia |

| | |
|---|---|
| | (short sighted) technically have a disability, but provided they have glasses or contact lenses do not necessarily require SEN support. |
| **SEN Support** | A less intensive form of SEND provision received by most pupils identified as having special educational needs or disabilities. It is school-led and school-funded and may include short-term interventions such as speech and language therapy, or additional support with reading and learning.[5] |
| **Education, Health and Care Plan (EHCP)** | A more intensive form of SEND provision for children and young people whose needs require legally enforceable support. EHCPs are assessed and partly delivered by the local authority and can remain in place until the age of 25.[5] An EHCP can be delivered in both mainstream and special schools, depending on the child's needs. Funding is primarily provided by the local authority where the child or young person resides, with additional contributions from the educational setting they attend (which may be located in a different local authority area). EHCPs replaced Statements of SEN following the 2014 education reforms.[39] |
| **SEND Code of Practice** | Statutory guidance published in January 2015 for identifying, assessing, and supporting children with SEND in England, used by schools, local authorities, and health services.[5] |
| **Graduated approach** | A step-by-step method used in schools to identify and support children with SEND, involving assess, plan, do, and review cycles recommended in the SEND code of practice.[40] |
| **Local Offer** | Information published by each local authority about the education, health, and social care services available for children and young people aged 0–25 with SEND. It explains what support is expected to be available locally and how to access it. The Local Offer must be clear, regularly updated, developed with input from families, and include services both within and outside the local area. It also provides guidance on eligibility, decision-making, support, and how to give feedback or make complaints. [41] |
| **Local Government & Social Care Ombudsman (LGSCO)** | An independent body that investigates complaints about local authorities, including issues related to education, social care, and SEND services. The Ombudsman investigates whether a council has acted unfairly or failed to follow proper procedures. The service is free, impartial, and usually used after all local complaint processes have been exhausted.[42] |
| **6. Sociodemographic variables** | |
| **6.1 Developmental indicators** | |
| **Gender** | NPD variable based on the legal sex of the pupil as reported by the parent or carer. It is recorded as either 'male' or 'female' and does not capture gender identity. [43] |
| **Sex at birth** | Based on sex recorded during HES birth admission(male/female) in analyses for question 1, and from NPD for analyses used in questions 2 and 3. |
| **Month of birth** | Refers to the month in which a child is born, as recorded in the HES birth admission. In the context of education in England |

| | |
|---|---|
| | (where the academic year runs from September to August) birth month has been shown to influence both cognitive and non-cognitive skill development, particularly in primary school.[44] Children born in summer months (e.g., July or August) may be nearly a year younger than some of their classmates born in autumn (e.g., September or October), placing them at a developmental disadvantage early in their schooling.[44] Month of birth was adjusted for in analyses by regrouping into four categories: September–November, December–February, March–May, and June–August. |
| **6.2 Health characteristics** | |
| **Birth weight** | Birth weight (in grams) was obtained from Hospital Episode Statistics (HES) birth admission records and supplemented using linked maternal health records where available. It was grouped into four categories: <2500g, 2500–4000g, >4000g, and missing.[45] |
| **Gestational age** | Gestational age (in completed weeks) was obtained from Hospital Episode Statistics (HES) birth admission records. It was used both as a continuous, week-by-week variable and grouped into categories for stratified analyses: very preterm (24–31 weeks), moderately preterm (32–33 weeks), late preterm (34–36 weeks), early term (37–38 weeks), and full term (39–41 weeks) and missing.[46] |
| **Phenotypes** | See Section 1of this table. |
| **6.3 Racial-ethnic characteristics** | |
| **Ethnicity** | NPD variable recording mode of ethnicity across any School Census that the pupil was included in. Can be recorded in the NPD as any of these categories: Asian, Chinese, Black, Mixed, White, Any Other Ethnic Group, and Unclassified/Missing. [43] |
| **Primary language** | NPD variable indicating pupil's primary language group. Recorded to be English, Other than English – which is often labelled 'English as an alternative language (EAL), and Unclassified/Missing.[43] |
| **6.4 Region/local government** | |
| **Local authority** | Local authorities (152 across England) are organisations that are officially responsible for a range of local services for individuals and businesses, including education, some health services and social services. Encompasses 24 county councils, 59 unitary authorities, 36 metropolitan district councils and 33 London boroughs.[47] |
| **Region of residence** | NPD variable taken from earliest recording in School Census (usually Reception or Year 1). Using the local authority of a pupil's recorded home address, we create groups corresponding to Government Office Region (GOR). North East, North West, Yorkshire and the Humber, East Midlands, West Midlands, East of England, London, South East, South West.[43] |
| **6.5 Indicators of social disadvantage** | |
| **Maternal age at delivery** | Maternal age at delivery was obtained from the HES birth admission record. A 'young mother' was defined as being under 20 years old at delivery and used as an indicator of social |

| | |
|---|---|
| | disadvantage. In stratified analyses, maternal age was categorised into six groups: <20, 20–24, 25–29, 30–34, 35–39, and 40+ years.[46] |
| **Index of Multiple Deprivation (IMD)** | The Index of Multiple Deprivation (IMD) is the official measure of relative deprivation in England. It ranks small geographic areas called Lower-layer Super Output Areas (LSOAs) from most to least deprived based on a combination of factors such as income deprivation, employment deprivation, education and crime.[48] |
| **Income Deprivation Affecting Children Index (IDACI)** | The Income Deprivation Affecting Children Index (IDACI) is a measure of the proportion of children aged 0–15 living in income-deprived households within a local area in England. It is a sub-domain of the Index of Multiple Deprivation (IMD) and is used to assess levels of child poverty.[49] In ECHILD, the IDACI decile of each child's lower super output area of residence is linked to their pupil record. |
| **Free school meal eligibility (FSM)** | A measure indicating whether a child meets the criteria to receive free school meals based on family income and receipt of certain benefits. Families need to register to be indicated as eligible in the NPD, thus children may not be recorded as such if they do not sign up. FSM eligibility is often used as a proxy for socioeconomic disadvantage in education data.[50] |
| **6.6 Outcomes used in ECHILD analyses** | |
| **Planned admissions** | A planned admission is a hospital admission where the decision to admit has been made in advance and the patient is admitted at a later date. These are typically part of a planned course of treatment or care, rather than in response to an urgent clinical need. Planned admissions usually follow from an outpatient consultation or a previous hospital episode and are scheduled to take place after a waiting period. Planned admissions were determined using if the *admimeth* variable in HES was coded as 11, 12, 13, 81, 84, 89.[36][51] |
| **Unplanned admissions** | An unplanned admission refers to a hospital admission that occurs unexpectedly and without prior scheduling. These admissions typically arise from an urgent or emergency clinical need. Unplanned admissions were determined using *admimeth* codes: 21, 22, 23, 24, 25, 28, 2A, 2B, 2D. [36] [51] |
| **Mortality** | The proportion of children in the cohort who died during the follow-up period, expressed relative to all live births in the cohort. Mortality is reported for two time windows:<br>i) Before school entry (from birth up to the start of Year 1);<br>ii) During primary school (from Year 1 to the end of Year 6). Used in Question 1, Paper 2. |
| **Attainment at KS1 and KS2** | See section 5 of table. |
| **Good Level of Development (GLD)** | See section 5 of table. |
| **Authorised absences/ unauthorised** | See section 5 of table. |

| | |
|---|---|
| **Total absences** | Total absences as a proportion of all half day periods available in the school year. Used in Questions 1, 3 (Papers 3, 16) |

#### Table S2: Abbreviations of commonly used terms in the HOPE study.

| **Abbreviation** | **Full term** |
|---|---|
| ADHD | Attention-Deficit/Hyperactivity Disorder |
| AP | Alternative provision |
| CA | Congenital Anomalies |
| CAMHS | Child and Adolescent Mental Health Services |
| CFA | Children and Families Act |
| CHC | Chronic Health Condition |
| CLP | Cleft Lip/ Palate |
| CP | Cerebral Palsy |
| CQC | Care Quality Commission |
| CYP | Children and Young People |
| DfE | Department for Education |
| DHSC | Department for Health and Social Care |
| ECHILD | Education and Child Health Insights from Linked Data |
| EHCP | Education, Health and Care Plan |
| EYFSP | Early Years Foundation Stage Profile |
| FGD | Focus Group Discussion |
| FSM | Free School Meals |
| GA | Gestational Age |
| GLD | Good Level of Development |
| HES | Hospital Episode Statistics |
| HES A&E | Hospital Episode Statistics Accident & Emergency |
| HES APC | Hospital Episode Statistics Admitted Patient Care |
| HOPE | Health Outcomes of young People throughout Education |
| IDACI | Income Deprivation Affecting Children Index |
| ICC | Intraclass Correlation Coefficient |
| IMD | Index of Multiple Deprivation |
| IV | Instrumental Variable |
| KS | Key Stage |
| LA | Local Authority |
| LGSCO | Local Government and Social Care Ombudsman |
| LO | Local Offer |
| LSOA | Lower-layer Super Output Area |
| MAT | Multi-Academy Trust |
| N1 | Nursery 1 |
| N2 | Nursery 2 |
| NC | National Curriculum |
| ND | Neurodisability |
| NHS | National Health Service |
| NIHR | National Institute of Health and Care Research |
| NPD | National Pupil Database |

| | |
|---|---|
| ONS | Office for National Statistics |
| PMR | Pupil Matching Reference |
| PPI | Public and Professional Involvement |
| PRU | Pupil Referral Unit |
| PSC | Programme Steering Committee |
| RCT | Randomised Control Trial |
| SAT | Single-Academy Trust |
| SEN | Special Educational Needs |
| SENCO | Special Educational Needs Coordinator |
| SEND | Special Educational Needs and Disability |
| SEND CoP | Special Educational Needs and Disability Code of Practice |
| TTE | Target Trial Emulation |
| VA | Voluntary Aided |
| VC | Voluntary Controlled |

#### Box 1: Summary of policy context for SEND provision in England

A shift from 2001 [37] towards inclusion of children with SEND in mainstream schooling was followed by the Children and Families Act (2014),[38] and the SEND Code of Practice (2015),[5] which aimed to improve SEND provision and social care services for vulnerable children and families. In 2022, a House of Lord's review of the 2014 Act concluded the government had 'ultimately failed' to improve outcomes, due partly to poor data collection across services and failure of implementation.[52] These legislative changes coincided with two further changes. Austerity-driven cuts to local authority (LA) budgets from 2010 reduced funding for SEND and social care support. Secondly, the expansion of more autonomous academy schools from 2007, contributed to uneven SEND provision across areas and schools.[53] [54] A national analysis using education data for primary schools in England reported that the primary school that a child attend makes more difference to whether they are assigned SEND provision than anything else.[55][56]

By age 16, one-third of all children in English state schools have been assigned SEND provision.[57] Approximately 30% of all children are assigned only SEN Support, which is decided and provided by schools. Between 3-5% of all children are assigned an Education, Health and Care Plan (EHCP), which is assessed, decided and part-funded by local authorities.[5] SEN Support includes in-class assistance, teaching aides or adaptive learning, while EHCPs provide tailored and personalised support for more complex needs. Half of the children who receive an EHCP attend a special school.[38][5] Since 2010, SEN Support reduced and, since 2016, use of more expensive EHCPs increased, leading to rising annual costs of SEND provision and exceeding £11 billion in England in 2025.[54][58]

The National Pupil Database records SEND provision each school term.[25] Based on consultation with users and providers of SEND services, we interpret this record to indicate assignment of SEND provision. It does not necessarily reflect receipt of provision. No measure of need for provision is routinely collated in the NPD. The HOPE study addressed this problem by using health phenotypes, such as coded information in hospital records to indicate neurodisability, congenital anomalies, chronic health conditions and gestational age at birth, along with developmental and social indicators, to proxy need for SEND provision.

### Appendix 2: Details of public and professional involvement.

#### Public and Professional Involvement (PPI)

PPI was supported by the teams from Cambridge (Ford/ Saxton) working in collaboration with the team from Exeter (Logan/ Boddy). We consulted young people and parent-carers using SEND services, and professionals with experience of delivering or commissioning services to advise the study. Collectively, we refer to these three groups as stakeholders. These consultations were separate from but informed the qualitative studies that involved stakeholders (young people, parent-carers and professionals) as participants in research. In a few cases, the same individuals were consulted as advisors and involved as participants in studies, or as peer interviewers.

##### 1. Involving young people and parent-carers – The HOPE Study

*Independent Programme Steering Committee*

Before the study began, we recruited two parent-carers (KE and JO) of children receiving SEND provision to be part of the independent Programme Steering Committee to shape the direction of the HOPE Study and provide strategic oversight of the study and its relevance to policy and practice.

*PPI advisory group members*

At the beginning of the study, we began recruiting members to join three PPI advisory groups through our steering committee members, wider professional networks, and by advertising on social media and through SEND journals. Group membership remained open throughout the study and the current numbers per group are as follows:

1) children and young people with SEND (n=14).

2) parents/carers of children with SEND (n=34).

In addition to having our own PPI groups, we consulted young people and parent-carers from two external advisory groups. One group, called FLARE (standing for Friendship, Learning, Achieve, Reach and Empower), involved children and young people with disability. The group was commissioned by the Department for Education and affiliated to the Council for Disabled Children. The other involved parent-carers of disabled children advising the Peninsula Children's Research Unit (PenCRU) and was affiliated to the University of Exeter and facilitated by Kate Boddy.

*How PPI members were involved*

The advisory groups were involved in several ways: online meetings, online drop-in sessions, and through the provision of written/emailed feedback. All PPI advisory members were reimbursed for their time in accordance with NIHR requirements. The PPI groups influenced The HOPE Study positively both to improve the quality of our research processes and tools, and to ensure the research itself was useful and relevant to service users. Below are three ways, with examples, of how the groups were involved and influenced the study:

1. **Ensuring questions asked in interviews and surveys were appropriate, relevant and likely to highlight factors influencing variation in SEND provision and outcomes**. For example, all PPI groups helped to develop our online survey. We received more than 200 question suggestions from the groups, and though we could not include all, we identified

common issues around trust between families and professionals, and training standards for SEND professionals, which were included in the survey.

2. **Review of language in documents.** We asked PPI group members to review our tools (such as topic guides) and our participant information sheets and consent forms to ensure the language was appropriate and understandable.
3. **Contribution as observers of focus group discussions (FGD):** We trained six groups pf parent-carers to observe focus group discussions with SEND professionals, to help us interpret the resulting data. Three of these parent-carers were able to attend six focus groups and provided input on the interpretation of the findings. These three also provided critical feedback on the report of the focus groups and were included as co-authors.

*How the PPI groups added to our conclusions, beyond data collection*

Many of the perspectives form advisory group members were echoed in the mixed methods research findings. Key messages, over and above what the mixed methods told us were:

1. There needs to be a radical overhaul of the education system to enable sufficient flexibility to accommodate and meet everyone's needs. The ambition for most children with SEN to be educated in mainstream settings and to reach their full potential will not be realised without policy change away from the narrow focus on core academic subjects; delivered by teachers with inadequate SEND training and resources; and where the emphasis on using exams to assess children's learning is experienced as punitive by many children. The latter is  particularly those with high levels of anxiety and who contend with sensory processing difficulties and sensory overload.
2. Young people with SEND who are at the point of leaving the education system, and for whom SEND will be lifelong, are encountering unfair barriers to employment and difficulties accessing adult health services due to differing thresholds for care in the post-16/post-18 period. The message from group members is that more attention needs to be paid to the 16-25 group with SEND to ensure they are supported and included in society.
3. The current SEND system is bringing many parents/carers to breaking point – and many have had to reduce hours or give up work completely to manage paperwork demands. It is clear that parent/carer advocacy is a critical factor in being able to access SEND services – too much depends on them at the moment. Our group members were also keenly aware that they are the people who have the resources (time, energy, money) to engage with what the system requires of them (albeit at a cost). Many parents/carers do not have capacity to do this, which is likely to mean that their children will have needs that remain unmet, exacerbating inequalities between groups. This is not about 'sharp elbowed parents' unfairly taking resources from others, rather that elbows should not need to be sharp to ensure basic provision.

#### 2. Involvement of professionals

*Independent Programme Steering Committee (PSC)*

The PSC was chaired by an expert in social education and health research, Professor Chris Bonell. Other members were Dr Karen Horridge, a senior community paediatrician, Dr Jo Hutchison, a researcher in SEND provision, and the two parent-carer members. The PSC met four times during the study and members were involved in additional discussions when their expertise was sought by the study team.

*Professional advisory group members*

The team at Cambridge University recruited a wider stakeholder group of professionals (*n*=35), mainly working in schools or health (e.g. therapists, psychologists) to deliver SEND provision, or in local authorities, social care or third sector SEND services.

### Appendix 3: ECHILD-derived study cohorts for quantitative analyses.

Table S3: List of ECHILD-derived cohorts used in HOPE sub-studies and in Figure S1.

| **Cohort Number** | **Cohort Name** | **Used in papers** |
|---|---|---|
| **1** | Neurodisability Birth Cohort | 2,10 |
| **2** | Neurodisability School Cohort (subset of cohort 1) | 2 |
| **3** | Neurodisability and Congenital Anomalies Outcomes Cohort | 3, 5, 7 |
| **4** | Gestational Age and Chronic Conditions Outcomes Cohort | 11 |
| **5** | Congenital Anomalies SEND Cohort | 9 |
| **6** | Inequalities in SEND Cohort – whole population | 15 |
| **7** | Local Authority Variation Cohort | 13 |
| **8** | Cerebral Palsy Sociodemographic Variation Cohort | 12 |
| **9** | Cerebral Palsy Cohort | 18 |
| **10** | Cleft Lip with Palate Cohort | 16 |

See Supplementary Figure 1 for cohort details.

Supplementary Figure 1. Timeline of birth years and follow-up years used in cohorts derived from ECHILD.

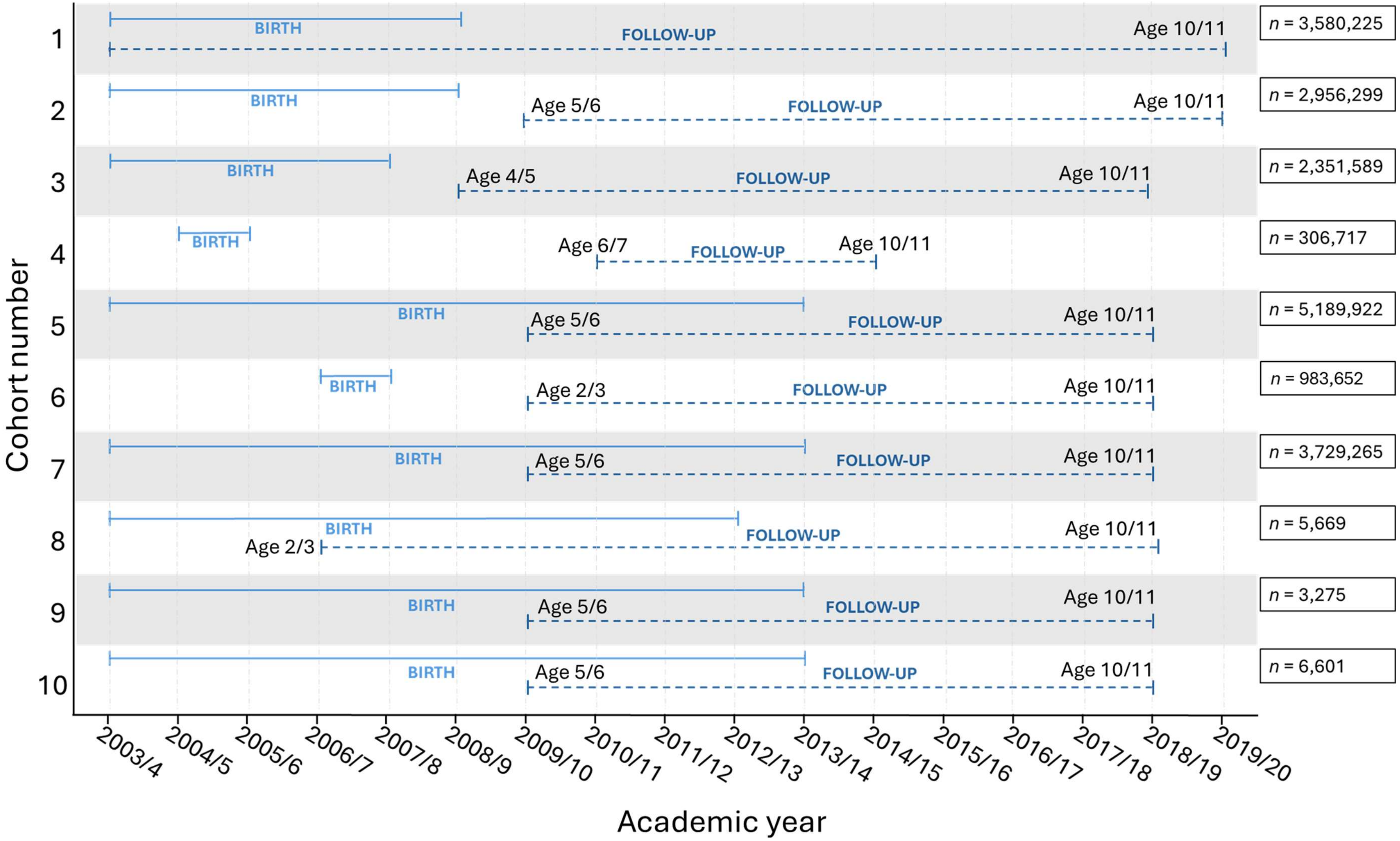

***Note**: Timeline ends in 2019/2020 due to disruptions caused by the COVID-19 pandemic which affected the availability and consistency of education and health records. Birth and follow-up year defined according to the academic calendar (e.g. 2003/4 includes 1st September 2003 to 31st August 2004, inclusive).*

### Appendix 4: List of HOPE sub-study publications.

| No. | Paper Name | Analysis cohort | Question (phenotype-colour coded) |
|---|---|---|---|
| **Research papers** 1 | Zylbersztejn A, Lewis K, Nguyen V, et al. Evaluation of variation in special educational needs provision and its impact on health and education using administrative records for England: umbrella protocol for a mixed-methods research programme. *BMJ Open*. 2023;13:e072531. Available from: https://doi.org/10.1136/bmjopen-2023-072531 | N/A | All Protocol |
| 2 | Zylbersztejn A, Rees P, D'Souza R, Logan S, Cant A, Gimeno L, et al. 2025;Phenotyping Neurodisability in Hospital Records in England: A National Birth Cohort Using Linked Administrative Data - Zylbersztejn - Paediatric and Perinatal Epidemiology - Wiley Online Library | 1,2 | Q1 & Q2a |
| 3 | Gimeno L, Zylbersztejn A, Cant A, Gilbert R, Harron K. Hospital admissions and school absences among primary school children with and without neurodisability: Linked administrative data cohort study in England. 2025; Under 2nd Review *DMCN* | 3 | Q1 |
| 4 | Gimeno L, Zylbersztejn A, Cant A, Gilbert R, Harron K. Planned and unplanned hospital admissions and health-related school absence rates in children with neurodisability: Protocol for a population-based study using linked education and hospital data from England. [version 1; peer review: 1 approved, 3 approved with reservations]. *NIHR Open Res* 2024; 4:26. Available from: https://doi.org/10.3310/nihropenres.13558.1 | 3 | Q1 protocol |
| 5 | Cant A, Zylbersztejn A, Gimeno L, Nguyen V, Tan J, Gilbert R, et al. Educational attainment among primary school children with neurodisability: A population-based cohort study using linked education and health data from England [Internet]. 2025 Jun. [cited 2025 23];2025.06.12.25329491. Available from: https://www.medrxiv.org/content/10.1101/2025.06.12.25329491v1 | 3 | Q1 |
| 6 | Cant A, Zylbersztejn A, Gimeno L, Nguyen V, Tan J, Gilbert R, et al. Primary school attainment outcomes in children with neurodisability: Protocol for a population-based cohort study using linked education and hospital data from England [version 1; peer review: 2 approved with reservations]. *NIHR Open Res* 2024, 4:28. Available from: https://doi.org/10.3310/nihropenres.13588.1 | 3 | Q1 protocol |
| 7 | Tan J, Cant A, Lewis KM, Nguyen V, Gimeno L, Zylbersztejn A, et al. Educational attainment of children with major congenital anomalies during primary school in England: a population cohort study using linked administrative data from ECHILD [Internet]. 2025 Jun. [cited 2025 24];2025.06.21.25329922. Available from: https://www.medrxiv.org/content/10.1101/2025.06.21.25329922v1 https://doi.org/10.1101/2025.06.21.25329922 | 3 | Q1 Congenital anomalies |
| 8 | Tan J, Cant A, Lewis KM, Nguyen V, Gimeno L, Zylbersztejn A, et al. Educational outcomes of children with major congenital anomalies: Study protocol for a population-based cohort study using linked hospital and education data from England [version 1; peer review: 2 approved, 2 approved with reservations]. *NIHR Open Res* 2024, 4:68. Available from: https://doi.org/10.3310/nihropenres.13750.1 | 3 | Q1 protocol |

| | | | |
|---|---|---|---|
| 9 | Peppa M, Lewis KM, De Stavola B, Hardelid P, Gilbert R. School-recorded special educational needs provision in children with major congenital anomalies: A linked administrative records study of births in England, 2003-2013. *Int J Popul Data Sci* 10:2519. https://doi.org/10.23889/ijpds.v10i1.2519 | 5 | Q1, Q2a & Q2d |
| 10 | Shumway J, Ellis J, Stephens A, De Stavola BL, Gilbert R, Zylbersztejn A. Hospital-recorded chronic health conditions in children with and without Down syndrome in England: a national cohort of births from 2003 to 2019. *Arch Dis Child* 2025 Mar.;110:221. https://doi.org/10.1136/archdischild-2024-327532 | 1 | Q1 |
| 11 | Libuy N, Gilbert R, Mc Grath-Lone L, Blackburn R, Etoori D, Harron K. Gestational age at birth, chronic conditions and school outcomes: a population-based data linkage study of children born in England. *International Journal of Epidemiology* 2023 Feb.;52:132–43. https://doi.org/10.1093/ije/dyac105 | 4 | Q2a<br>Week of gestation at birth |
| 12 | Lewis KM, Nguyen VG, Ní Chobhthaigh S et al. Sociodemographic variation in cumulative probability of recorded special educational needs and disability provision during primary school in England: A staggered cohort study of children with cerebral palsy [version 1; peer review: awaiting peer review]. *NIHR Open Res* 2025, 5:61. https://doi.org/10.3310/nihropenres.14006.1 | 8 | Q2b |
| 13 | Kate Lewis, Vincent Nguyen, Ruth Gilbert, Bianca De Stavola, Lorraine Dearden. Local authority variation in school-recorded special educational needs provision in Year 1 among children born in England, 2003-2013. Eur J Pub Health (in press July 2025) | 7 | Q2c |
| 14 | Lewis K, Nguyen V, Zylbersztejn A, Gilbert R, De Stavola B, Dearden L. Local authority variation in primary school-recorded special educational needs provision among children with major congenital anomalies: A research protocol. *NIHR Open Res* 2023 Oct.;3:50. https://doi.org/10.3310/nihropenres.13466.1 | 5 | Q2c<br>protocol |
| 15 | Bianca L De Stavola,[1*] Kate M. Lewis,[1] Vincent G. Nguyen,[1] Andrea Aparicio Castro,[1] Lorraine Dearden,[2] Ruth Gilbert,[1] on behalf of the HOPE study team. Is there evidence of inequalities in the provision, level, and timing of SEND in English primary schools? Preprint Is there evidence of inequality in the provision, level, and timing of SEND provision in English primary schools? \| medRxiv | 6 | Q2abc<br>Whole population |
| 16 | Nguyen V, Lewis K, Gilbert R, De Stavola B, Dearden L. The impact of early special educational needs provision on later hospital admissions, school absence and education attainment: A target trial emulation study of children with isolated cleft lip and/or palate. *PLoS One* 2025;20:e0327720. https://doi.org/10.1371/journal.pone.0327720 | 10 | Q3 |
| 17 | Nguyen VG, Lewis KM, Gilbert R et al. Early special educational needs provision and its impact on unplanned hospital utilisation and school absences in children with isolated cleft lip and/or palate: a demonstration target trial emulation study protocol using ECHILD [version 1; peer review: 1 approved, 2 approved with reservations]. *NIHR Open Res* 2023, 3:54 (https://doi.org/10.3310/nihropenres.13472.1) | 10 | Q3<br>protocol |
| 18 | Vincent Nguyen, Kate Lewis, Ruth Gilbert, Bianca De Stavola[1*], Lorraine Dearden. Early special educational needs provision and health and educational outcomes during primary school in children with cerebral palsy. *NIHR Open Res* 2025, **5**:65 (https://doi.org/10.3310/nihropenres.14015.1 | 9 | Q3 |
| 19 | Andrea Aparicio Castro, Bianca De Stavola, Lorraine Dearden. Assessing the short- and long-term effects of Special Education Needs (SEN) provision on unauthorised absences: a methodological comparison of causal contrasts and estimation methods. (for submission end of September 2025, plus technical report). | 6 | Q3 |

| | | | |
|---|---|---|---|
| 20 | Lorraine Dearden, Andrea Aparicio Castro, Kate M. Lewis,Vincent G. Nguyen, Bianca De Stavola The causal impact of special educational needs on education and health outcomes in England: a practitioner's guide. (for submission September 2025) | 6 | Q3 |
| 21 | Gains, H., Winterburn, I., Saxton, J., Matthews, J., Farr, W., Black-Hawkins, K. (2025). Identification, assessment and provision of special education needs in England: a cross-sectional survey to compare perceptions of young people, parents/carers and professionals. European Journal of Special Needs Education, 1–17. https://doi.org/10.1080/08856257.2025.2491192 | | Q4a-c Quantitative online survey |
| 22 | Albajara Saenz, A., Winterburn, I., Matthews, J., Burn, A. M., Ford, T & Saxton, J. Lessons from Local Area SEND Inspections: A Content Analysis of Ofsted Outcome Letters. British Educational Research Journal (under review). | | 4a-e Mixed methods |
| 23 | Jacob Matthews, Isaac Winterburn, Jennifer Saxton, Nazneem Nazeer, Eleanor Chatburn, Ariadna Albajara Sáenz, Miyuki Komachi, William Farr, Kristine Black-Hawkins, Tamsin Ford. Exploring experiences of the English special educational needs system through an online survey of young people, and parents and carers. British Journal of Special Educational Needs https://nasenjournals.onlinelibrary.wiley.com/doi/full/10.1111/1467-8578.70043 | | 4a-c Quantitative online survey |
| 24 | William Farr, Isaac Winterburn, Jacob Matthews, Jennifer Saxton, Tamsin Ford. Surveying the Professional Experience of Special Educational Needs Provision in England. British Journal of Special Education (under review). | | 4a-c Quantitative online survey |
| 25 | Saxton, J., Matthews, J., Winterburn, I., Casey, H., Barnes, S., Zylbersztejn, A., Hall, P., Tripp, C., Black-Hawkins, K. and Ford, T. Exploring the experiences and outcomes of children and young people receiving support for special educational needs over time in England: a qualitative study. In Frontiers in Education (Vol. 10, p. 1564583). Frontiers. doi: 10.3389/feduc.2025.1564583 | | 4a-f Interviews with children with SEND |
| 26 | Saxton, J., Winterburn, I., Matthews, J., Anderson, J., Farr, W., Minhas, S., Smith, S., Ford, T. Right support, right place, right time; right mess!' Professionals' views on factors influencing the SEND system and outcomes for children and families in England: A qualitative study. Archives of Public Health (under review). | | 4a-f Focus groups with SEND professionals |
| 27 | Saxton, J., Albajara Saenz, A., Williams, O., Matthews, J., Winterburn, I., Chatburn, E., Nazeer, N., Black-Hawkins, K., Ford, T. Examining local level variation in Special Educational Needs and Disabilities (SEND) service provision and associated data sources in England: A scoping review. Humanities, Social Science & Communication. (under review) | | 4a-f Scoping review |
| 28 | Saxton, J., Barnes, S., Gains, H., Chatburn, E., Ford, T. Annual trends in LGSCO complaints and outcomes relating to Special Educational Needs and Disabilities in England: 2018 to 2023. Research Papers in Education (under review) | | 4b-f Quantitative analysis |
| 29 | Saxton, J., Burn, A.M., Zhang, X., Toulmin, H., Parker, J., Casey, H., Matthews, I., Tripp, C., Barnes, S., Hall, P., Black-Hawkins, K., Gains, H., Ford, T. Barriers, enablers and outcomes reported by parents engaged with the special educational needs system in England: A qualitative study. PlosOne (under review) | | 4a-f Interviews with parent-carers |
| 30 | Matthews, J., Black-Hawkins, K., Basu, A., Necula, A-I., Downs, J., Ford, T. & Saxton, J.; (2024). To what extent do England's local offer websites adhere to the statutory guidance as set out in the special | | 4a-c & e Mixed methods analysis |

| | | | |
|---|---|---|---|
| | educational needs and disabilities code of practice? British Educational Research Journal, 50, 1724–1740. https://doi.org/10.1002/berj.3996 | | |
| **Blogs** 31 | Jacob Matthews, Kristine Black-Hawkins, Arina Basu, Andreea-Ioana Necula, Jonny Downs, Tamsin Ford, Jennifer Saxton. Research shows “critical information gaps” in most SEND local offer websites, including EHCP eligibility criteria. https://www.specialneedsjungle.com/research-critical-information-gaps-send-local-offer-websites-ehcp-eligibility-criteria/ | | Blog based on analysis of Local Offer for SEND websites |
| 32 | Ruth Gilbert, Kate Lewis, Will Farr. Special educational needs policy requires research infrastructure. BERJ Jan 2025. https://www.bera.ac.uk/blog/special-educational-needs-policy-requires-research-infrastructure | | Policy blog |
| 33 | Tamsin Ford & Jennifer Saxton, December 2022. Our three rights-based wishes for reforming SEND services in England. https://www.place2be.org.uk/about-us/news-and-blogs/2022/december/our-three-rights-based-wishes-for-reforming-send-services-in-england/ | | Blog for users of services |
| **Key finding flyers** 34 | Sarah Barnes, Jacob Matthews, Isaac Winterburn, Jennifer Saxon, Kristine Black-Hawkins, Tamsin Ford, December 2022. Key findings: Children and Young People’s Survey https://www.ucl.ac.uk/population-health-sciences/sites/population_health_sciences/files/hope_study_-_cyp_survey_key_findings_flyer.pdf | | Layperson / visual summary |
| 35 | Sarah Barnes, Jacob Matthews, Isaac Winterburn, Jennifer Saxon, Kristine Black-Hawkins, Tamsin Ford , December 2022. Key findings: Parents and Carer’s Survey https://www.ucl.ac.uk/population-health-sciences/sites/population_health_sciences/files/hope_study_-_parent_carer_survey_key_findings_flyer.pdf | | Layperson / visual summary |
| 36 | Sarah Barnes, Jacob Matthews, Isaac Winterburn, Jennifer Saxon, Kristine Black-Hawkins, Tamsin Ford, December 2022. Key findings: SEND Professional’s Surve https://www.ucl.ac.uk/population-health-sciences/sites/population_health_sciences/files/hope_study_-_wider_stakeholder_group_survey_key_findings_flyer.pdf | | Layperson / visual summary |
| 37 | Sarah Barnes, Jacob Matthews, Isaac Winterburn, Jennifer Saxon, Kristine Black-Hawkins, Tamsin Ford, December 2022. Key findings Survey Summaries https://www.ucl.ac.uk/population-health-sciences/sites/population_health_sciences/files/hope_study_-_national_surveys_key_findings_poster.pdf | | Layperson / visual summary |
| 38 Code | Code used for analyses can be found referenced in papers or below:<br>1. In the ECHILD code list repository https://code.echild.ac.uk/<br>2. In the UCL Child Health Informatics Group repository for the HOPE study eg: https://github.com/UCL-CHIG/HOPE_neurodisability<br>3. By contacting authors | | Code used in analyses |

### Appendix 5: Methods and key findings on what service experiences and policies tell us about the underlying process of SEND provision

Research question 4: summaries of sub-study findings mapped to questions 4 a to f on: a) SEND identification, b) assessment, c) provision, d) outcomes, e) local service capability/functioning and f) national policy context

1. **In Appendix 5, 'children' is used to refer to children and young people aged 13 to 25 years and 'parents' is used to refer to parents and carers.**

| Sub-study | Method | Initial identification (4a) | Assessments and planning (4b) | Provision (4c) | Outcomes (4d) | Local service capability (4e) | National policy context (4f) |
|---|---|---|---|---|---|---|---|
| **Qualitative studies** | | | | | | | |
| Right support, right place, right time; right mess!' Professionals' views on factors influencing the SEND system and outcomes for children and families in England: A qualitative study (under review).[59]<br><br>Appendix 4 paper no.26 | Focus-group discussions (N=6) with 35 SEND professionals, using a topic guide co-developed with parents of children with SEND. | • Initial identification of additional needs is improving in the classroom, especially with well trained, experienced SENCOs.<br>• Complexity of SEND is increasing.<br>• There are increasing numbers of children presenting to services with SEND.<br>• Quieter children with unmet needs may be overlooked in favour of disruptive children.<br>• Some professionals view the increase in SEND presentations as due to a general worsening of social circumstances, whilst others think we are better at recognising conditions. | • Interagency working is seriously impaired, which causes delays to assessments and diagnoses.<br>• Poor information sharing systems, low capacity to put together coordinated responses, poor working relationships, logistical problems of meeting, lack of specialists, and lack of shared understanding of what a child's needs are. Therefore, lots of referrals 'bounce back' which harms family's trust in professionals.<br>• There are insufficient numbers of trained professionals to implement the system properly. | • Support is often delayed.<br>• Lack of support in school or inappropriate educational placements are common and harmful for children.<br>• SEND provision depends on parent understanding of the system, advocacy ability and financial resources.<br>• Trust and good relationships with children and families are important for SEND provision, but are usually fraught, characterised by "mistrust", and worsen in the "limbo period" of waiting.<br>• The system should much earlier prepare young people with SEND for adult life, higher education and employment.<br>• Professionals are being taken away from preventive work to deal with EHCP assessment and plan related paperwork.<br>• There are ongoing disputes between professionals and LAs, and families and LAs about provision. | • Long-term harm to parent and child mental health from delays and navigating the system.<br>• Families face out of pocket expenditure pursuing diagnoses and gathering evidence for tribunals.<br>• Children from families who cannot advocate are missing from the system, worsening outcomes and increasing inequalities in health and education.<br>• Unmet needs mean that some children never have a chance to feel successful and be proud of their achievements.<br>• Relationships between professionals and families, and between professionals are often very poor. | • Services are patchy and inconsistent because there is growing demand and more bureaucracy than ever before, with fewer SEND professionals and less money available to meet requirements.<br>• Information-sharing systems, standardisation of documents and processes, and interagency meetings improve quality/consistency of SEND activities. | • SEND provision operates within a broader system of national education which is not suitable for some children (e.g. curriculum demands, focus on exams, narrow subject focus, mainstream classroom environments).<br>• Other policies such as forced academisation, and stringent school attendance expectations can be indirectly and directly harmful for children with SEND.<br>• Many professionals are being expected to task shift (e.g. teachers and mental health provision) to compensate for England-wide gaps/delays in CAMHS services.<br>• There are too few educational psychologists for the SEND system to work and preventive services and early interventions to take place.<br>• SEND-related discrimination in the employment sector |

| | | | | | | | |
|---|---|---|---|---|---|---|---|
| | | | | • Decisions based on funding rather than need. | | | needs to be addressed. |
| Barriers, enablers and outcomes reported by parents engaged with the special educational needs system in England: A qualitative study (under review).[60]<br><br>Appendix 4 paper no.29 | One-to-one interviews with N=22 parents. Drawn life 'timelines' were used to gain a comprehensive picture of participants' experiences of engaging with the system over time. | • Children with SEND are often 'unseen and unheard', misunderstood by school staff and mislabelled as naughty which could delay identification – particularly autism and ADHD, as well as children with communication difficulties. | • Several parents reported negative experiences of CAMHS assessments.<br>• Some parents reported unreasonably high thresholds to qualify for provision.<br>• Parents felt like they were ping pong balls in terms of referrals bouncing between departments. | • Legal protections and advocacy ability of parents and professionals are key enablers of SEND provision but come at a huge cost to families and legal processes can be very slow.<br>• Several parents reported premature discharge from CAMHS when children were still in crisis.<br>• Poor relationships and communication between professionals and with parents were common and undermined provision quality, caused delays, and made the system hostile.<br>• Decisions about provision were often based on poorly written and/or outdated plans.<br>• Parents excluded from multi-agency decisions which appeared to be against best interests of child. | • Positive outcomes for some: health, education and social, autonomy, independence.<br>• Negative outcomes for others: lost years of education and opportunities within settings, damage to parent and child mental health from delays and treatment by SEND professionals (particularly LAs), lack of aspiration for children by professionals also worsens their long-term prospects, parents losing out financially and in careers because of poor provision.<br>• Parents who cannot 'fight' may disengage – worsening inequalities. | • Inconsistent definitions of SEND at LA level (e.g. dyslexia).<br>• Variable processes at LA level in terms of when you can apply for ECH assessments.<br>• Variable thresholds for care/eligibility. | • Educational policies lack long-term vision for children with SEND.<br>• Lack of safety nets within health and social care for when CYP reach adulthood and parents can no longer provide intensive support. |
| Exploring the experiences and outcomes of children and young people receiving support for special educational needs over time in England: a qualitative study [61]<br><br>Appendix 4 paper no.25 | One-to-one interviews (with 15 children and young people aged 13-25, with SEND. A timeline approach was used in conjunction with a semi-structured interview schedule. | • Late identification due to low awareness of some conditions (e.g. autism in girls, which may present differently to boys).<br>• Most children wanted to be identified and supported much earlier.<br>• Many children and young people said they were not listened to about their needs. | • Many children spent years on waiting lists for CAMHS and for diagnoses, often unsupported in the interim. | • Primary schools were described by some as being able to provide good needs-led support.<br>• Some SENCOs stood out as being able to offer effective tailored provision, whilst others were criticised.<br>• Some schools would not offer support without a diagnosis.<br>• Parent and child advocacy ability were critical for getting provision in place. | • Sensory overload was commonly reported, and provision that enabled children to manage this was effective for improving educational and mental health outcomes.<br>• Time spent waiting for assessments and interventions was harmful for education and mental health.<br>• Many children reported positive effects of provision on their independence. | • Lack of local specialist SEND and CAMHS professionals were causing delays to provision and worsening the outcomes of children.<br>• School resources sometimes dictated the quality of provision (e.g. physical space for sensory room). | • Many children said teachers would benefit from additional neurodiversity training.<br>• Many children said exams were an unfair format for them to demonstrate their skills. |

| | | | | | | | |
|---|---|---|---|---|---|---|---|
| | | | | • Some provision was inappropriate and ineffective. | • Exam-support and mentoring was helpful.<br>• Transitions that were not well managed (e.g. primary to secondary) could exacerbate problems.<br>• Effectiveness of provision was strongly influenced by school context, ethos and senior leadership.<br>• Individually tailored provision was more helpful than generic. | | |
| **Sub-study** | **Method** | **Initial identification (4a)** | **Assessments and planning (4b)** | **Provision (4c)** | **Outcomes (4d)** | **Local service capability (4e)** | **National policy context (4f)** |
| **Online surveys of children and young people, parents/carers and SEND professionals** | | | | | | | |
| Identification, Assessment and Provision of Special Education Needs in England: A cross-sectional survey to compare perceptions of young people, parents/carers and professionals. [62]<br><br>Appendix 4 paper no.21 | Self-report, cross-sectional surveys completed by young people with SEND (N = 77), parents (N = 770) and SEND professionals (N = 863). | • Parents rated their perception of identification services as positive (N=214, 28%), neutral (N = 145, 19%) and negative (53%, N = 409).<br>• SEND professionals rated their perception of providing services for identification as positive (N=296, 34%), neutral (N = 237, 27%) and negative (N = 243, 28%).<br>• Most parents disagreed that professionals had enough training in SEND identification.<br>• Most SEND professionals reported they did have appropriate training to identify children and young people who may have SEND. | • Few parents agreed that enough information was provided to them about their child's additional needs during assessment from teaching staff (26% N = 192), health services (32%, N=226), or the LA (16%, N = 108).<br>• Few parents agreed that enough information was provided directly to their child from education staff (N = 176, 24%), health services (N = 178, 26%), or the LA (N = 78, 12%).<br>• 45% of professionals (N=390) disagreed that the assessment process adequately supported families who were less able to advocate for themselves. | • Experiences of provision were split between negative and positive.<br>• 49% (N = 374) of parents rated their overall perception of provision as negative or neutral (N = 201, 19%), whilst 25% (N = 193) reported it to be positive.<br>• Most children agreed that they "would have liked extra support for my learning earlier in my education' (N = 57, 74%) and fewer endorsed neutral (N = 7, 9%) or disagreed with the statement (N = 8, 10%).<br>• Perceptions of SEND provision were mixed for SEND professionals; 45% (N = 389) rated provision as positive, neutral (N = 242, 28%) and negative (22%, N = 187). | • This paper does not report on outcomes. | • Most children (N = 55, 71%) have not heard about the LO for SEND.<br>• The LO website was the least common source of information for parents (N = 8, 1%). Instead, parents used online forums (N = 165, 21%), social media (N = 145, 19%), independent advocates (N = 135, 18%) and other parents (N = 123, 16%) for ongoing information gathering around their child's SEN.<br>• Most parents (72%, N = 551) disagreed that there are enough support services available to parents of children with SEN.<br>• Most SEND professionals had heard of the LO (83%, N = 715). However, only 39% (N = 336) agreed their LO provides all the information it is supposed to, and 23% (N = 197) agreed their LA was able to provide all the SEND services it was supposed to. | • Whilst professionals rate their training in SEN identification as adequate, parents have negative views of professionals' level of knowledge and skills. This suggests that training might not always be effectively translated into practice.<br>• Parents say there is inadequate support for families who are less able to advocate for themselves and SEND provision depends on financial/social capital.<br>• Professionals say that young people are not adequately involved in their assessments at present, despite this being a statutory expectation. |

| | | | | | | | |
|---|---|---|---|---|---|---|---|
| Exploring the experiences of the English Special Educational Needs System through an online survey of young people, and parents and carers[63]<br><br>Appendix 4 paper no.23 | Self-report, cross sectional survey of young people with SEND (N=77) and parents/carers (N=770) | • Only a third of parents agreed that education, health and local authorities are supportive in identifying SEN.<br>• The three main barriers that parents reported to identification are long waiting lists, lack of understanding in education settings and lack of specialist SEND professional in their LA.<br>• Most (91%) children reported that their parents talk to them about their need for extra support, whereas only 51% agreed the same of teachers and health professionals (45%).<br>• In total, 47% of children were identified as needing extra support in pre-school (25%) or Key Stage One (22%). | • 80% of children trusted their parents to make the right decisions about their extra support needs, while only 42% trusted their teachers and 48% trusted health professionals.<br>• This indicates potential vulnerability for children without strong parental support.<br>• 48% of children felt involved in decision making during their assessment process.<br>• Children described their experience of assessment as positive (40%), frustrating (32%), and stressful (25%). Words describing negative experiences were more commonly selected. | • Satisfaction for support provided is low among parents: 41% agree that their child's EHCP responds to all their needs and 34% agree that their child's EHCP or SEN statement matches what their child receives.<br>• 87% of children reported that their parents listened to what they said about their learning needs, whilst 55% agreed the same of teachers.<br>• 62% of children reported that their teacher included them in lessons.<br>• 54% of children believed their extra support helped them access the same classes as their peers, demonstrating the importance of timely and accurate provision. | • Survey responses suggest that children EHCPs lack information about future goals and aspirations and are often not appropriately ambitious.<br>• 35% of parents agreed that their child's EHCP or SEN statement includes appropriately ambitious learning outcomes.<br>• 49% of parents agreed that future goals and aspirations are included in their child's EHCP or SEN statement. | • A minority of parents reported that key service providers had a good or very good understanding of what support their youngest child with SEND required: education settings (26%), health services (20%), and local authorities (14%).<br>• Most parents reported negative experiences when liaising with LAs about SEND provision.<br>• 38% of parents had paid for private assessment, suggesting lack of availability or satisfaction with the public sector assessments. | • No relevant data. |
| Surveying the Professional Experience of Special Educational Needs Provision in England (under review).[64]<br><br>Appendix 4 paper no. 24 | Self-report, cross sectional survey of SEND professionals (N=770). | • A minority (38%) of SEND professionals agreed that there is effective communication between agencies involved in SEND identification.<br>• There is variation between professionals from different sectors (e.g. in their self-reported knowledge and confidence).<br>• 46% of LA staff agreed that there is effective communication during identification, followed by 44% of health professionals and 36% of education staff.<br>• Most professionals felt confident in identifying children with SEND (86%). | • This study does not report on formal assessments and plans. | • Most professionals (63%) felt neutral or disagreed that they can provide sufficient support to families in their LA most of the time.<br>• Half of all professionals (50%) felt confident most of the time in designing and providing services for children. This was consistent across LA, health, education and other professionals.<br>• 71% of professionals felt confident in communicating with families about SEND provision, with higher agreement from LA professionals (85%) and lowest agreement from healthcare (63%). | • This study does not report on outcomes. | • Only 20% of professionals felt they had sufficient time and resources to deliver good quality SEND for all children with SEND in their LA.<br>• Over 95% over professionals reported at least one barrier to providing good quality SEND.<br>• The most frequently selected barriers were lack of LA funding (20%), length of waiting lists (18%), insufficient access to SEND specialists (14%) and excessive caseload (10%).<br>• The least selected barriers were burnout (1%), communication with parents (1%), poor senior leadership (1%), relationships | • There were overall mixed reports about whether professionals had sufficient training to design and deliver good quality SEND provision in the LA where they work(ed) most of the time.<br>• Over half (58%) agreed that they do have sufficient training.<br>• Views on having sufficient training were similar across education (60%), LA professionals (60%) and health (54%). However, only 44% of other professionals agreed. |

| | | | | | | | |
|---|---|---|---|---|---|---|---|
| | | Confidence was highest amongst health professionals (91%) followed by LA professionals (88%), education professionals (85%) and other professionals (79%). | | | | with Social Care professionals (1%), difficulties interpreting and applying the SEND CODE of practice (1%), relationships with PCs (0.8%) and relationships with education professionals (0.5%). | |
| **Sub-study** | **Method** | **Initial identification (4a)** | **Assessment and planning (4b)** | **Provision (4c)** | **Outcomes (4d)** | **Local service capability (4e)** | **National policy context (4f)** |
| **Document review** | | | | | | | |
| Examining local level variation in Special Educational Needs and Disabilities (SEND) service provision and associated data sources in England: A scoping review (under review).[65]<br><br>Appendix 4 paper no.27 | A scoping review that identified N=18 peer reviewed studies, N=105 grey literature studies and compiled open access data sources including SEND data at LA or Multi-academy Trust level. | • Inconsistent and inequitable identification of SEND within a single MAT.<br>• Inconsistent identification of autism at LA level, from different phenotypic prevalence and/or 'differences in detection or referral'.<br>• Role of the SENCO is crucial to identification and onward referrals, but scale and capacity to carry out work is hugely variable - wide array of tasks, expectations of multiple stakeholders & differing seniority. | • Quality of EHCP processes and documents were undermined in multiple ways.<br>• EHCPs not always needs-based, influenced by need type (e.g. SEMH harder to validate), parental advocacy, SENCO experience, interagency working, inconsistent application of the law.<br>• Outsourcing of EHCP writing affects quality and co-production with families. 'Copy-paste' plans & written outcomes often not functional or SMART, lack of tailoring.<br>• Inconsistent methods being used and barriers to meaningfully capturing child voice/preference in EHCPs.<br>• Inspections found that excessive bureaucracy was increasing waiting times. | • 'Gap between ideology and implementation' of CFA, in context of no additional staff or funding.<br>• TAs are most confident, effective, and can apply specialist training when they are linked with LA specialist teams.<br>• Preparation for adulthood programmes should start earlier (by 13-14 years) and the offer should be broadened beyond education settings to promote belonging and create mentoring and employment opportunities.<br>• Clear interagency communication is required to ensure successful transition from child to adult health services.<br>• Post-16 options provided by LAs are too limited. Parents wanted an impartial list of options, and not to be steered by LAs to inappropriate or funding-based placements, and LAs to listen to children's preferences.<br>• Large variation at LA level about awareness of the LO | • Negative outcomes for parents: complaints & redressal data shows high parent distress from engaging with statutory processes (delays, protracted process, unmet needs and expectations, fear for the future for children).<br>• Financial outcomes for LAs: Mediation (not about placement) may save LAs money overall for less complex cases as can avoid more costly tribunals (low quality evidence).<br>• Positive outcomes from post-16 schemes when LAs are involved in apprenticeships and internships as it allows effective targeting of underserved groups.<br>• Educational outcomes: Children with SEND are disproportionately excluded, and there is no way appeal or influence the new placement that is named; in parallel LAs have low understanding about alternative provision locally, affecting the quality of commissioning and placement decisions. | • Staff burnout, inconsistent processes, poor interagency working, and low-quality LOs.<br>• Funding cuts and limited training and resources locally drive increasing local variation in provision.<br>• High local variation in the % of EHCPs, mediation and appeals against LA decisions relative to child population.<br>• Increasing demand for SEND services, which many LAs were not ready for.<br>• Feedback by service users about the LO for SEND (all from low quality research) suggests between-LA variation in people's satisfaction with it the corresponding website (the extent it was useable, with accurate and up to date information).<br>• There is a wealth of unused administrative data that would enable meaningful reporting on local variation in SEND identification and provision, including disaggregating by socio-demographic factors to examine equity of service coverage and use. | • Professionals need 'clear guidelines and systematic, standardised training' to improve plans & implementation.<br>• Switch from Independent Appeal to Independent Review Panels, and government drive to convert schools to academies negatively affected children who were excluded from school (e.g. LAs less able to challenge schools about decisions).<br>• Academies often affected by reporting delays about newly identified children with SEND compared to previous system.<br>• SEND CoP guidance led to fewer children being recorded in SEN registers, in turn resulting in fewer specialist teachers in some LAs. |

| | | | | | | | |
|---|---|---|---|---|---|---|---|
| | | | | of SEND provision and uptake of related services such as short breaks/respite care. | | | |
| To what extent do England's local offer (LO) websites adhere to the statutory guidance as set out in the special educational needs and disabilities code of practice [66]<br><br>Appendix 4 paper no.30 | Secondary content analysis of LO websites. Where possible analysis was also conducted on annual reports, policy documents, surveys and meeting minutes provided on LO websites. | • LO websites are a potentially crucial first information source for people in the early stages of noticing their child's additional needs - but there are information gaps on LO websites, which contravene current SEND legislation.<br>• LO websites commonly lack accessibility features, failing to meet the needs of many service users. This may exacerbate sociodemographic inequalities by forming a barrier to information access. | • Across England, 89% of LO websites provide information about the EHCP process. However, only 47% of LAs provide eligibility criteria in clear, simple, plain language with bullet points or checklists. The language used on LO websites needs to be simplified to facilitate engagement of children and parents. | • There is high variation between LAs in the quality of LO websites. This reinforces the concern that SEND provision depends on a postcode lottery.<br>• Information on available health services is provided widely on LO websites, with all LAs providing information on services related to mental health.<br>• All LAs provided information on available schools, however only 73% included a transport policy statement.<br>• Only 31% of LO websites signpost to offline resources or alternative means for those with limited internet access. | • This paper does not report on outcomes. | • There is wide variation in the quality of LO websites, with some very poor examples and other websites that offer excellent content and navigability.<br>• There is evidence that many LAs included children and parents in developing the LO website. However, only 8% complied with the Equality Act when preparing the LO.<br>• Whilst 99% of LAs provide information about the option of having a personal budget, only 64% describe the services for which these budgets can be used. | • There is limited evidence for effective regional partnerships or information sharing, demonstrated by high variability of LO websites both within and between regions.<br>• The CoP states 'must' criteria as legal requirements. However, there is greater adherence to 'should' criteria, which are not legal requirements. Many LO websites require substantial updates to reach legal standards. |
| Lessons from Local Area SEND Inspections: A Content Analysis of Ofsted Outcome Letters (under review).[67]<br><br>Appendix 4 paper no.22 | Secondary content analysis of Ofsted/ CQC inspection letters (N=36 initial inspection letters and N=34 revisit letters). | • Most common strengths inspectors reported about SEND identification: Inter-agency working, health services/support and interventions, accurate, timely and early identification of needs.<br>• Common weaknesses also included health services, and accuracy of needs identification, as well as educational provision and support at school. | • Widespread problems with EHCP process often meaning the LA failed their inspection, and sometimes their reinspection.<br>• EHCP issues reported concerned: quality, timeliness, outcomes, accuracy, poor multi-agency input, lack of monitoring, leading to inappropriate provision. | • Most common strengths inspectors reported about meeting needs were inter-agency working, health services/support and interventions, and relationships with parents and carers.<br>• Common weaknesses were also relationships with parents, health services, as well as educational provision and support at school and EHCPs. | • Most common outcomes improved were educational/academic, employment and training, and educational provision and support at schools.<br>• Most common outcomes needing improvement: Leaders' awareness and system utilisation and educational/ academic outcomes and progress. | • Overlap in common strengths and weaknesses indicates variation between LAs.<br>• Most common area of significant weakness triggering a written statement of action by LAs were: LA leaders and leadership (91.2% of letters), SEND provision and support, and EHCPs (both in 82.4% of letters).<br>• In revisits following a written statement of action (87.1%) were found to make sufficient progress improving leadership, (82.4%) improved SEND support and (82.4%) improved EHCPs. | • Our data provide benchmarks for current practices at LA level and identify common areas of strength and concern that can be monitored in new iterations of the inspection framework being rolled out across England. |

| | | | | | | | |
|---|---|---|---|---|---|---|---|
| Annual trends in Local Government and Social Care Ombudsman complaints and outcomes relating to Special Educational Needs and Disabilities in England: 2018 to 2023 (under review).[68]<br><br>Appendix 4 paper no.28 | Secondary analysis of LGSCO administrative data, 2018 to 2023. Descriptive statistics were generated to explore variation in SEND-related complaints to the ombudsman in England by year, complaint type, and subsequent outcomes (upheld, not upheld, not considered). | • This paper does not report on initial identification. | • Complaints regarding the assessment and reviews of needs fall within the "SEN" complaints category and this was the most frequent type of complaint across all data years.<br>• The rate of SEN related complaints (complaints per 10,000 pupils with EHCPs or with 'SEN support') are increasing and accounting for an increasingly large proportion of complaints relative to other causes. | • Absolute numbers of complaints in all categories (alternative provision, transport, COVID, school admissions, school exclusions, SEN, and other) have risen steadily since 2021.<br>• The complaint rate (complaints per 10,000 pupils with EHCPs or with 'SEN support') increased more than four-fold, from 1.42 to 6.12. | • At least 80% of SEN and alternative provision complaints have been upheld in the favour of parents over time.<br>• School transport complaints were increasingly upheld over time (stabilising at around 75%).<br>• There were no complaints within the LGSCO remit about school exclusions. | • There is considerable variation in complaint rate across LAs.<br>• Over time there is an increasing complaint rate in an increasing number of LAs.<br>• Over time, there are fewer LAs receiving 0 complaints a year.<br>• LGSCO SEN complaints category reflects complaints on the provision of EHCP plans, transitions, budgets and direct payments. It is therefore not possible to isolate whether the assessment of children's SENs is driving the increase in complaints in this category. | • LGSCO complaints categories include alternative provision, COVID-19, school admissions, school exclusions, school transport, SEN and other complaints.<br>• COVID-19 complaints upheld have increased over the data years from 71% in 2020/21 to 88% in 2022/23.<br>• Other complaint types show slight decreases in 2020/2021, which may be because the LGSCO did not consider complaints for a 3-month period (February to April 2021) due to COVID-19 restrictions. |

### Appendix 6: RECORD Checklist.

**The RECORD statement – checklist of items, extended from the STROBE statement, that should be reported in observational studies using routinely collected health data.**

| | Item No. | STROBE items | Location in manuscript where items are reported | RECORD items | Location in manuscript where items are reported |
|---|---|---|---|---|---|
| **Title and abstract** | | | | | |
| | 1 | (a) Indicate the study's design with a commonly used term in the title or the abstract (b) Provide in the abstract an informative and balanced summary of what was done and what was found | | RECORD 1.1: The type of data used should be specified in the title or abstract. When possible, the name of the databases used should be included.<br><br>RECORD 1.2: If applicable, the geographic region and timeframe within which the study took place should be reported in the title or abstract.<br><br>RECORD 1.3: If linkage between databases was conducted for the study, this should be clearly stated in the title or abstract. | 1.1 Title and Abstract – objectives refer to ECHILD database<br><br>1.2 England in title and abstract. Timeframe in abstract<br><br>1.3 In Title and Abstract |
| **Introduction** | | | | | |
| Background rationale | 2 | Explain the scientific background and rationale for the investigation being reported | | | Pages 1,2 |
| Objectives | 3 | State specific objectives, including any prespecified hypotheses | | | Page 3<br><br>Hypotheses for causal methods on page 8 |

| Methods | | | | | |
|---|---|---|---|---|---|
| Study Design | 4 | Present key elements of study design early in the paper | | | Shown for each question in methods pages 6-9 |
| Setting | 5 | Describe the setting, locations, and relevant dates, including periods of recruitment, exposure, follow-up, and data collection | | | Page 4 and Appendix 1 Box 1. |
| Participants | 6 | *(a) Cohort study* - Give the eligibility criteria, and the sources and methods of selection of participants. Describe methods of follow-up<br><br>*Case-control study* - Give the eligibility criteria, and the sources and methods of case ascertainment and control selection. Give the rationale for the choice of cases and controls<br><br>*Cross-sectional study* - Give the eligibility criteria, and the sources and methods of selection of participants<br><br>*(b) Cohort study* - For matched studies, give matching criteria and number of exposed and unexposed | | RECORD 6.1: The methods of study population selection (such as codes or algorithms used to identify subjects) should be listed in detail. If this is not possible, an explanation should be provided.<br><br>RECORD 6.2: Any validation studies of the codes or algorithms used to select the population should be referenced. If validation was conducted for this study and not published elsewhere, detailed methods and results should be provided.<br><br>RECORD 6.3: If the study involved linkage of databases, consider use of a flow diagram or other graphical display to demonstrate the data linkage process, including the number | 6.1 Cohort derivation shown in Appendix 3, Table S3 and Fig1.<br><br>Further details referred to in substudies listedfor cohorts in Table S3.<br><br>6.2, Phenotyping studies and algorithms for populations reported in substudy papers 2 to 10 are summarised on page 6 and listed in Appendix 4.<br><br>6,3 Flow diagrams are not shown but are available in substudy reports papers 2-10 |

| | | | | | |
|---|---|---|---|---|---|
| | | *Case-control study* - For matched studies, give matching criteria and the number of controls per case | | of individuals with linked data at each stage. | |
| Variables | 7 | Clearly define all outcomes, exposures, predictors, potential confounders, and effect modifiers. Give diagnostic criteria, if applicable. | | RECORD 7.1: A complete list of codes and algorithms used to classify exposures, outcomes, confounders, and effect modifiers should be provided. If these cannot be reported, an explanation should be provided. | Variables are described in Appendix 1, Table S1. |
| Data sources/ measurement | 8 | For each variable of interest, give sources of data and details of methods of assessment (measurement).<br><br>Describe comparability of assessment methods if there is more than one group | | | Variable sources in Appendix 1, Table S1 and in substudy papers. |
| Bias | 9 | Describe any efforts to address potential sources of bias | | | Addressing bias is intrinsic to the TTE framework applied to descriptive (Qu1,2a-d) and causal questions (Qu3).<br><br>Efforts are summarised in methods (pages 6-9)<br><br>Details are referenced in substudies and listed in Appendix 4. |

| | | | | | |
|---|---|---|---|---|---|
| Study size | 10 | Explain how the study size was arrived at | | | Summarised for substudies in Appendix 3, S3 Table and Figure 1; Further details referenced in substudies. |
| Quantitative variables | 11 | Explain how quantitative variables were handled in the analyses. If applicable, describe which groupings were chosen, and why | | | Summarised in in methods pages 6-9, variable descriptions in Appendix 1, and results (pages 10-18). Full explanations referenced in substudies (Appendix 4). |
| Statistical methods | 12 | (a) Describe all statistical methods, including those used to control for confounding<br>(b) Describe any methods used to examine subgroups and interactions<br>(c) Explain how missing data were addressed<br>(d) *Cohort study* - If applicable, explain how loss to follow-up was addressed<br>*Case-control study* - If applicable, explain how matching of cases and controls was addressed<br>*Cross-sectional study* - If applicable, describe analytical methods taking account of sampling strategy | | | a) Summarised in methods and results<br>b) Addressed by different substudies eg:<br>Qu 2b and 2c, separate analyses of the whole population (cohort 6) and children with cerebral palsy (cohort 8; page 7,8,13-15)<br>Qu 3; cleft lip with palate and cerebral palsy (page 8,9,18 and papers 16-18 (Appendix 4)<br>c) Missing data addressed only in substudy papers.<br>d) Only in substudy papers |

| | | | | | |
|---|---|---|---|---|---|
| | | (e) Describe any sensitivity analyses | | | |
| Data access and cleaning methods | | .. | | RECORD 12.1: Authors should describe the extent to which the investigators had access to the database population used to create the study population.<br><br>RECORD 12.2: Authors should provide information on the data cleaning methods used in the study. | Described in substudies specific to each cohort (referenced and listed in Appendix 4; also see Appendix 3 for cohort – substudy links). |
| Linkage | | .. | | RECORD 12.3: State whether the study included person-level, institutional-level, or other data linkage across two or more databases. The methods of linkage and methods of linkage quality evaluation should be provided. | Summarised in methods: Questions 1, 2 a to d, and Question 3 (pages 6-9) |
| **Results** | | | | | |
| Participants | 13 | (a) Report the numbers of individuals at each stage of the study (*e.g.*, numbers potentially eligible, examined for eligibility, confirmed eligible, included in the study, completing follow-up, and analysed)<br><br>(b) Give reasons for non-participation at each stage. | | RECORD 13.1: Describe in detail the selection of the persons included in the study (*i.e.*, study population selection) including filtering based on data quality, data availability and linkage. The selection of included persons can be described in the text and/or by means of the study flow diagram. | Summarised in Appendix 3 (Table and Figure) |

| | | | | | |
|---|---|---|---|---|---|
| | | (c) Consider use of a flow diagram | | | |
| Descriptive data | 14 | (a) Give characteristics of study participants (*e.g.*, demographic, clinical, social) and information on exposures and potential confounders<br><br>(b) Indicate the number of participants with missing data for each variable of interest<br><br>(c) *Cohort study* - summarise follow-up time (*e.g.*, average and total amount) | | | Not shown but referred to in substudy papers (via references and Appendix 4). |
| Outcome data | 15 | *Cohort study* - Report numbers of outcome events or summary measures over time<br><br>*Case-control study* - Report numbers in each exposure category, or summary measures of exposure<br><br>*Cross-sectional study* - Report numbers of outcome events or summary measures | | | Shown graphically in results (pages 11-18) and substudies referenced for details (Appendix 4) |
| Main results | 16 | (a) Give unadjusted estimates and, if applicable, confounder-adjusted estimates and their precision (e.g., 95% confidence interval). Make clear which | | | Main results are summarised and presented graphically. Details are referenced in substudy papers (Appendix 4).<br><br>Unadjusted and adjusted results are shown |

| | | | | | |
|---|---|---|---|---|---|
| | | confounders were adjusted for and why they were included<br><br>(b) Report category boundaries when continuous variables were categorized<br><br>(c) If relevant, consider translating estimates of relative risk into absolute risk for a meaningful time period | | | graphically for question 2c only.<br><br>Confounders are listed in methods for questions 2abc and 3 (pages 6-9) |
| Other analyses | 17 | Report other analyses done—e.g., analyses of subgroups and interactions, and sensitivity analyses | | | Relevant substudy papers referred to in results to questions 1, 2a-d and 3 (pages 10-13) |
| **Discussion** | | | | | |
| Key results | 18 | Summarise key results with reference to study objectives | | | Page 21 |
| Limitations | 19 | Discuss limitations of the study, taking into account sources of potential bias or imprecision. Discuss both direction and magnitude of any potential bias | | RECORD 19.1: Discuss the implications of using data that were not created or collected to answer the specific research question(s). Include discussion of misclassification bias, unmeasured confounding, missing data, and changing eligibility over time, as they pertain to the study being reported. | Page 22 |
| Interpretation | 20 | Give a cautious overall interpretation of results considering objectives, limitations, | | | For question 3, pages 17-18, and in substudies (Appendix 4). |

| | | | | | |
|---|---|---|---|---|---|
| | | multiplicity of analyses, results from similar studies, and other relevant evidence | | | |
| Generalisability | 21 | Discuss the generalisability (external validity) of the study results | | | Pages 21-23 |
| **Other Information** | | | | | |
| Funding | 22 | Give the source of funding and the role of the funders for the present study and, if applicable, for the original study on which the present article is based | | | Page 24 |
| Accessibility of protocol, raw data, and programming code | | .. | | RECORD 22.1: Authors should provide information on how to access any supplemental information such as the study protocol, raw data, or programming code. | Access to data on page 24.<br>Protocols, detailed substudy papers and access to code listed in Appendix 4. Link to full reports, published and underreview provided for reviewers. |

*Reference: Benchimol EI, Smeeth L, Guttmann A, Harron K, Moher D, Petersen I, Sørensen HT, von Elm E, Langan SM, the RECORD Working Committee. The REporting of studies Conducted using Observational Routinely-collected health Data (RECORD) Statement. *PLoS Medicine* 2015; in press.

*Checklist is protected under Creative Commons Attribution (CC BY) license.

25. Jay MA, McGrath-Lone L, Gilbert R. Data Resource: the National Pupil Database (NPD). Int J Popul Data Sci 4:1101. https://doi.org/10.23889/ijpds.v4i1.1101

31. Early years foundation stage profile handbook.

32. Roberts N. Assessment and testing in primary education in England. 2025 Jul. [cited 2025 30]; Available from: https://commonslibrary.parliament.uk/research-briefings/cbp-7980/

33. Understanding scaled scores at key stage 1 [Internet]. GOV.UK 2025 May [cited 2025 30]; Available from: https://www.gov.uk/guidance/understanding-scaled-scores-at-key-stage-1

34. Understanding scaled scores at key stage 2 [Internet]. GOV.UK 2025 Jul. [cited 2025 30]; Available from: https://www.gov.uk/guidance/understanding-scaled-scores-at-key-stage-2

35. School attendance and absence [Internet]. childlawadvice.org.uk [cited 2025 30]; Available from: https://childlawadvice.org.uk/information-pages/school-attendance-and-absence/

36. Gimeno L, Zylbersztejn A, Cant A, Gilbert R, Harron K. Hospital admissions and school absences among primary school children with and without neurodisability: Linked administrative data cohort study in England. 2025;

50. Department for Education. Free school meals. Guidance for local authorities, maintained schools, academies and free schools. UK Assets Publishing Service: Department for Education; 2025.

51. National Health Service (NHS). Elective Admission Type - NHS Data Dictionary Archive [Internet]. 2024; Available from: https://archive.datadictionary.nhs.uk/DD%20Release%20May%202024/attributes/elective_admission_type.html

52. Children and Families Act 2014: A failure of implementation.

53. Ogden K, Phillips D. How have English councils' funding and spending changed? 2010 to 2024. 2024 Jun. [cited 2025 23]; Available from: https://ifs.org.uk/publications/how-have-english-councils-funding-and-spending-changed-2010-2024. https://doi.org/10.1920/re.ifs.2024.0318

54. Sibieta L, Snape D. Spending on special educational needs in England: something has to change. 2024 Dec. [cited 2025 5]; Available from: https://ifs.org.uk/publications/spending-special-educational-needs-england-something-has-change. https://doi.org/10.1920/re.ifs.2024.0949

55. Jo Hutchinson. Identifying pupils with special educational needs and disabilities [Internet]. London: Education Policy Institute; 2021 [cited 2025 23]. Available from: https://epi.org.uk/publications-and-research/identifying-send/
